# Associations of longitudinal BMI percentile classification patterns in early childhood with neighborhood-level social determinants of health

**DOI:** 10.1101/2023.06.08.23291145

**Authors:** Mehak Gupta, Thao-Ly T. Phan, Félice Lê-Scherban, Daniel Eckrich, H. Timothy Bunnell, Rahmatollah Beheshti

## Abstract

**Background:** Understanding social determinants of health (SDOH) that may be risk factors for childhood obesity is important to developing targeted interventions to prevent obesity. Prior studies have examined these risk factors, mostly examining obesity as a static outcome variable.

**Methods:** We extracted EHR data from 2012-2019 for a children’s health system that includes 2 hospitals and wide network of outpatient clinics spanning 5 East Coast states in the US. Using data-driven and algorithmic clustering, we have identified distinct BMI-percentile classification groups in children from 0 to 7 years of age. We used two separate algorithmic clustering methods to confirm the robustness of the identified clusters. We used multinomial logistic regression to examine the associations between clusters and 27 neighborhood SDOHs and compared positive and negative SDOH characteristics separately.

**Results:** From the cohort of 36,910 children, five BMI-percentile classification groups emerged: always having obesity (n=429; 1.16%), overweight most of the time (n=15,006; 40.65%), increasing BMI-percentile (n=9,060; 24.54%), decreasing BMI-percentile (n=5,058; 13.70%), and always normal weight (n=7,357; 19.89%). Compared to children in the decreasing BMI-percentile and always normal weight groups, children in the other three groups were more likely to live in neighborhoods with higher poverty, unemployment, crowded households, single-parent households, and lower preschool enrollment.

**Conclusions:** Neighborhood-level SDOH factors have significant associations with children’s BMI-percentile classification and changes in classification. This highlights the need to develop tailored obesity interventions for different groups to address the barriers faced by communities that can impact the weight and health of children living within them.

**Impact Statement:** This study demonstrates the association between longitudinal BMI-percentile patterns and SDOH in early childhood. Five distinct clusters with different BMI-percentile trajectories are found and a strong association between these clusters and SDOH is observed. Our findings highlight the importance of targeted prevention and treatment interventions based on children’s SDOH.

## 1. Introduction

Childhood obesity is a complex multifactorial disease.^1–3^ The ecological model of childhood obesity^4^ points to the important role of not only individual-level but also neighborhood-level SDOH in the development of childhood obesity. Some of the main neighborhood-level SDOH that have been linked to childhood obesity include single-parent households, housing insecurity, lower socioeconomic-status, lower parental educational-status, and employment.^5–14^ Historically under-resourced populations affected by the negative SDOH are not only disproportionately burdened by childhood obesity but have also shown a lower success rate of obesity prevention interventions.^15^ Targeting these community-level risk factors can help provide children with a healthy environment to prevent obesity.

Interventions designed to prioritize at-risk communities could be impactful in early childhood as most childhood and adult obesity begins early in life – as early as age two.^16^ Other important time-points for weight gain (e.g., adiposity rebound) occur before age five and definitely by around age seven,.^17,18^ Baidal et al^16^ found that modifiable risk factors like family socioeconomic-status (SES) and childcare attendance were associated with childhood obesity in the first two years of life, and Tester et al^19^ found that poverty and single-parent household status were associated with severity of obesity in children age 2-5.

There is a wealth of literature describing the relationship between selected SDOH factors and childhood obesity using cross-sectional analyses.^20–34^ However, these cross-sectional studies assume that all children with obesity are a homogeneous group with similar growth patterns, but the obesity growth pattern of children while developing obesity could have different implications for intervention.^35^ For example, children who have obesity from birth may have genetic and epigenetic risk factors and intervention may need to occur in the perinatal period, whereas children born with normal weight but then develop obesity during early childhood may benefit from interventions that can impact their environment and lifestyle. Therefore, to better understand the impact of SDOH on a child’s risk of obesity at different time-periods, it would be important to identify how SDOHs are associated with different weight trajectories of children throughout early childhood.^35,36^

While many existing longitudinal studies evaluate BMI, BMI-z, or BMI-percentile trajectories in children,^37–39^ with many also describing how these trajectories are associated with SDOHs.^40–50^ However, there have been no studies to date that have applied advanced algorithmic clustering techniques to establish distinct groups of patients with clinically-relevant trajectories (e.g., normal weight to obesity as defined by CDC) and few studies that have examined these associations in early childhood or weight trajectories with ungrouped neighborhood-level SDOH variables to better understand the individual contribution of SDOH which can better inform policy changes to target factors.^39,44,51^ Therefore, in this work, we aim to study the association between clinically-relevant BMI-percentile status throughout early childhood and a series of major neighborhood-level SDOH and individual-level socio-demographics using an electronic health record (EHR) dataset.

## 2. Materials and Methods

### 2.1 Data source

We obtained patient-level EHR data from Nemours Children’s Health - a large, multi-state pediatric health system in the US. Our study was approved by the Nemours Institutional Review Board. The initial dataset was extracted from PEDSnet dataset of children 0-21 years old with at least 5 years of medical data with patient records from 2002-19 for all inpatient and outpatient visits.^52^ Patients with complex medical conditions (requiring intensive treatment, hospitalizations, etc.) of type 1 diabetes, cancer, or sickle cell diseases (full list in Supplemental-S8) which are normally excluded from our data center’s extractions are also excluded from this cohort.

### 2.2 Study sample and anthropometric measurements

Due to regular pediatric visits in early childhood, our original data had a higher number of weight and height measurements among younger children (who have at least one visit between 0 to 7 years), with fewer measurements documented with increasing age and the largest drop in measurements after age seven. Because of this and because of our focus on early childhood, we included only 67,242 children of age 0-7. Because our original dataset mostly comprised young children, we only excluded 787 children in this step. We excluded 11 patients with no weight or height measurements or demographic data and 19,052 without a minimum of 5 years of recorded weight and height measurements. Since weight and height measurements for children were recorded over irregular intervals, we segmented the time-series data into non-overlapping 3-month windows to obtain smooth time-series with equal intervals. For every window, we took the median weight and height measurements. If weight and height measurements were missing in a certain 3-month window, we mark it as missing. We excluded 7,286 children with more than 80% missing rate for recorded weight and height measurements. Here we note that out of 26,338 (19,052+7,286) patients excluded in the previous two steps 15% lived in a census with more than 20% population with annual income below the federal poverty level compared to 23% in our final cohort. This shows that our final cohort had enough representation of the low and high SES populations compared to the excluded population to fulfill our study objective.

We classify patients based on Centers for Disease Control and Prevention (CDC) categories that use BMI-percentile, as having a normal weight (BMI-percentile *<* 85), overweight (85 *≤* BMI-percentile *<* 95), or obesity (BMI-percentile *≥* 95). Weight and height were used to calculate BMI and reference BMI-percentile provided by the World Health Organization (WHO)^53^ and CDC^54^ for children between 0 to 2 and older than 2 years, respectively. We choose to use BMI-percentile to remain consistent with the measurements used for the cohort across 0-7 years of age.^55–58^ Additional notes about our choice of BMI-percentile are presented in Supplemental-S1.

Finally, to keep the focus of our study on studying SDOH associations between children with normal, overweight, or obesity BMI-percentiles, we also excluded underweight patients (BMI-percentile *≤* 5) and never crossed the threshold of 85 (more details in Supplemental-S2 and Table-S1). This resulted in a cohort of 36,910 patients. All cohort selection steps are summarized in Figure 1 and Table 1 shows the general characteristics of our final cohort.

**Table 1:** Characteristics of the full cohort.

|  |  |
| --- | --- |
| **Population (N)** | 36,910 |
| **Male/Female** | 19,920/16,990 |
| **Mean BMI-percentile (±STD)** | 64 (±7.18) |
| **Ethnicity:** |  |
| Hispanic | 5,076(13.7%) |
| Non-Hispanic | 31,635(85.70%) |
| **Race:** |  |
| Asian | 638(1.72%) |
| Black | 15,195(41.16%) |
| White | 16,816(45.55%) |
| Other Race | 4,602(12.46%) |
| **Insurance:** |  |
| Public | 21,431(58.06%) |
| Private | 15,415(41.76%) |

**Figure 1:**
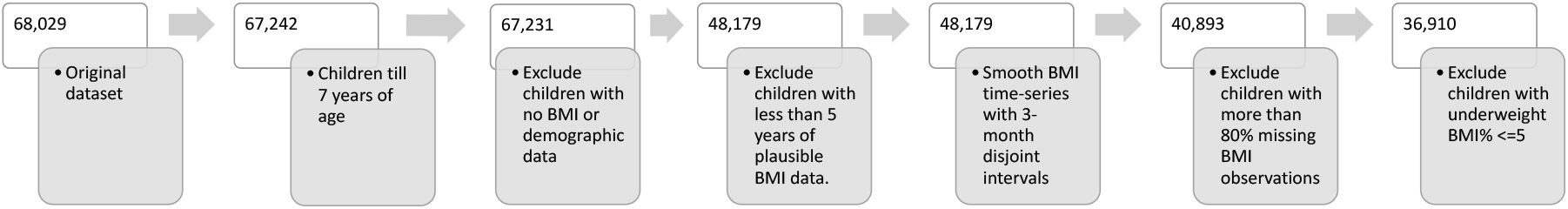
Cohort selection and processing steps. The initial extracted data of 68,029 children from Nemours Pediatric EHR only included children who had at least 5 years of medical data irrespective of how many observations in the 5 years. Please note that 5 years of medical data does not assure 5 years of weight and height measurements. Therefore, we exclude 19,052 children in step 3. Since young children tend to have more regular visits, our original dataset was mostly comprised of young children. Therefore, our dataset ends up having more children who have at least one visit between 0 to 7 years. Therefore, in step 1 only 787 children are excluded.

### 2.3 Linking patient data to neighborhood-level SDOH Data

Data collected in the EHR about patient race, ethnicity, and insurance type, and addresses were included in the cohort. The Census-Geocoder^59^ address look-up tool allowed us to map patient visits to 4,237 unique census tract geocodes. We mapped data obtained before 2010 to 2000 census tract boundaries and after 2010 to 2010 census tract boundaries. 23% of patients had a relocation, out of which 58% had no relocation after 3 years of age showing that their latest census-tract is also the longest-stayed census-tract. We used the latest census-tract for each patient.

To capture the most essential neighborhood-level SDOH that may influence child development we identified the union of variables from three standardized indexes: Child Opportunity Index (COI),^60^ Social Vulnerability Index (SVI),^61^ and Social Deprivation Index (SDI).^62^ We grouped these variables into five categories: 1) socioeconomic-status, 2) neighborhood household composition, 3) housing and transportation, 4) education and occupation, and 5) RUCA (rural–urban commuting area) code.^63^ Evaluating the association between individual SDOHs instead of high-level indexes and BMI-percentile trajectories allows us to determine which aspects of neighborhood risk have the most impact. This allows us to better inform practice and policy. All SDOHs are the ratio of the population belonging to that SDOH except for median household-income. RUCA is the ratio of the population with RUCA*≥*4. Additional details are presented in Supplemental-S3.

### 2.4 Clustering BMI-percentile trajectories

#### 2.4.1 Data-driven clustering

We first used a traditional data-driven clustering approach (i.e., using only raw data without further analysis) to find any homogeneous groups whose BMI-percentile category remained in the same category (normal, overweight, and obesity) over time. We found two groups: 1) BMI-percentile always in normal weight range and 2) BMI-percentile always in the obesity range. The remaining group had significant heterogeneity. The data-driven approach is method-independent to cluster the data for simple stratification.

#### 2.4.2 Algorithmic clustering

However, relying only on raw data for clustering generally offers limited power when working with complex data. An algorithmic approach can capture temporal and non-linear patterns in the data effectively. Therefore, we used a partitional clustering approach with dynamic time warping (DTW) distance to identify additional homogeneous clusters among the heterogeneous clusters. Technical details about our partitional clustering approach are presented in Supplemental-S4. Our results from this approach yielded three groups: over-weight, increasing, and decreasing BMI-percentile.

To further confirm the robustness of the three identified clusters in the choice of algorithmic method, we also used a separate clustering algorithm, latent class growth mixture modeling (LCGMM), which has been used in studying BMI trajectories.^64–67^ Technical details about LCGMM experiments and results are presented in Supplemental-S5.

### 2.5 Statistical Analysis

First, we compare the social characteristics between clusters and then compare the differences in positive and negative SDOH characteristics using analysis of variance (ANOVA) tests. We use multinomial logistic regression to analyze the association between the cluster’s membership and SDOH characteristics. We study the influence of each SDOH on the cluster membership individually. To do that, we dummy coded each categorical variable (Non-Hispanic, Hispanic, Asian, White, Black, Some Other Race, Private Insurance, and Public Insurance) to 0/1 and used the default reference category of 0 for each categorical variable.

## 3. Results

### Number of clusters

Using the data-driven clustering technique described in Section 2.4.1, we found two homogeneous clusters ‘always having obesity’ and ‘always having normal weight,’ leaving all remaining children in a heterogeneous pool of those with BMI-percentile recorded from at least two of the normal, overweight, or obesity categories. Next, by applying the algorithmic technique described in Section 2.4.2 on the heterogeneous pool, we identified three new clusters, which we can label as ‘overweight most of the time’, ‘increasing BMI-percentile’, and ‘decreasing BMI-percentile.’ For brevity, we refer to these five clusters as ‘obesity,’ ‘normal,’ ‘overweight,’ ‘increasing,’ and ‘decreasing.’ From our cohort of 36,910 children, 429 (1.16%) fell into the ‘obesity’ group, 15,006 (40.65%) fell into the ‘overweight’ group, 9,060 (24.54%) fell into the ‘increasing’ group, 5,058 (13.70%) fell into the ‘decreasing’ group, and 7,357 (19.89%) fell into the ‘normal’ group. All cluster trajectories are shown in Figure 2. Technical details about LCGMM experiments and results are presented in Supplemental-S5.

**Figure 2:**
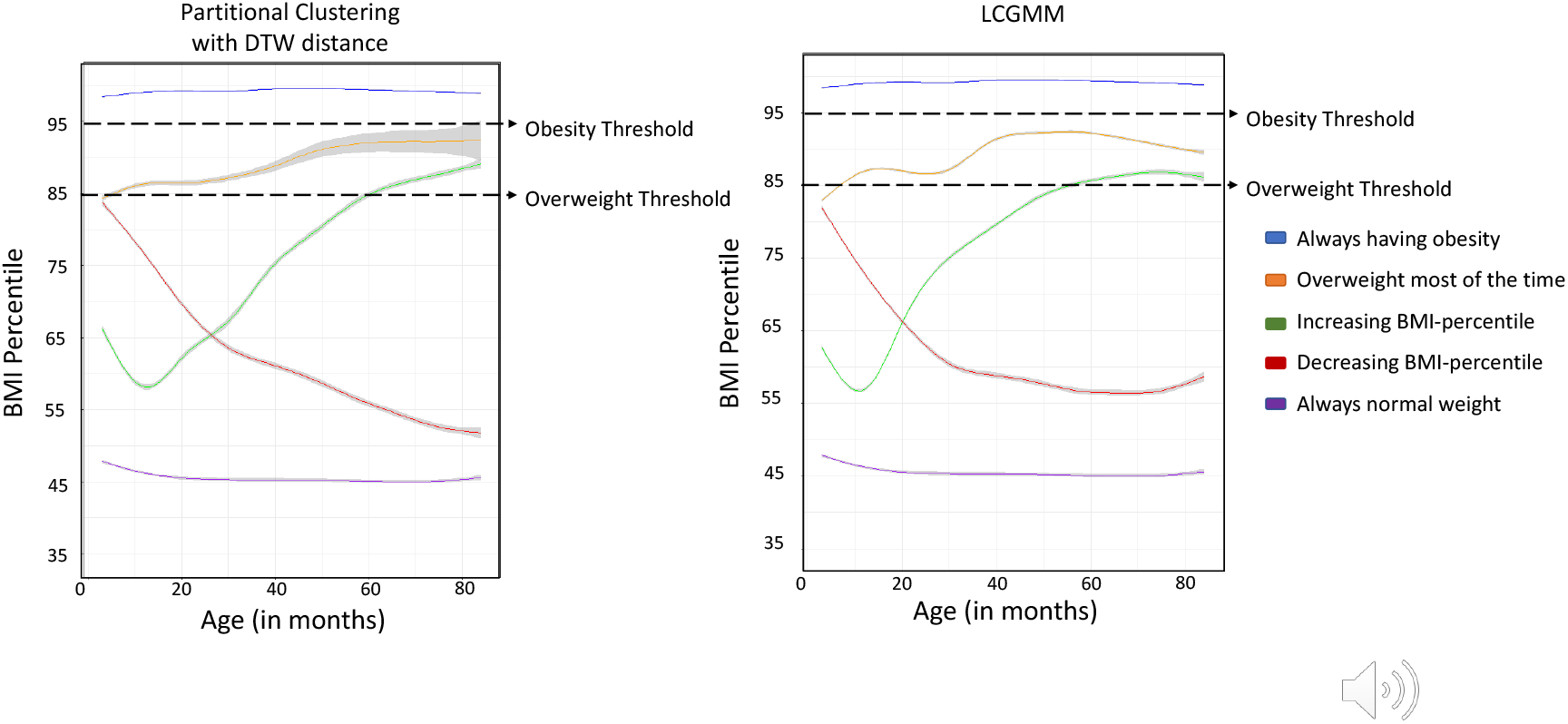
Five groups were identified in our study based on BMI-percentile trajectories of children between 0 to 7 years. We used a smoothing function (`geom_smooth` package in R) to represent each cluster by a smooth solid line with 95% CI. The figure on the left shows the trajectories found using data-driven clustering and partitional clustering with DTW. The figure on the right shows the trajectories found using data-driven clustering and partitional clustering with LCGMM. Technical details about LCGMM experiments and results are presented in Supplemental-S5.

### Race, ethnicity, insurance, and BMI-percentile of the five groups

Table 2 shows the proportion of children in each cluster, based on race, ethnicity, and insurance status. Groups indicating higher concerns for obesity (‘obesity’, ‘overweight’, and ‘increasing’) had a higher prevalence of Black (37% to 41%) and Hispanic (12% to 25%) children compared to other groups with less than 40% Black and less than 12% Hispanic. They also had a larger proportion with public insurance than private insurance. Whereas the ‘normal’ group had a greater than 50% White children and almost the same proportion with the public (51.92%) and private insurance (47.89%).

**Table 2:** Characteristics of five BMI-percentile trajectory clusters.

|  | Obesity<br>(Always having obesity) | Overweight<br>(Overweight most of the time) | Increasing<br>(Increasing BMI-percentile) | Decreasing<br>(Decreasing BMI-percentile) | Normal<br>(Always normal weight) | p-value<sup>a</sup> |
| --- | --- | --- | --- | --- | --- | --- |
| **Population [n(%)]** | 429(1.16%) | 15,006(40.65%) | 9,060(24.54%) | 5,058(13.70%) | 7,357(19.89%) | ** |
| **Male/Female** | 263/166 | 8,291/6,715 | 4,649/4,411 | 2,953/2,105 | 3,764/3,593 | ** |
| **Mean BMI-percentile (±SD)** | 99.10 (±1.06) | 93.26 (±21.37) | 65.14 (±26.54) | 58.45 (±24.52) | 41.18 (±20.40) | ** |
| **Ethnicity:** |  |  |  |  |  |  |
| Hispanic [n(%)] | 106(24.7%) | 2,470(16.46%) | 1,142(12.60%) | 620(12.25) | 738(10.03%) | * |
| Non-Hispanic [n(%)] | 321(74.28%) | 12,536(83.53%) | 7,792(86.%) | 4,412(87.22%) | 6,574(89.35%) | ** |
| **Race:** |  |  |  |  |  |  |
| Asian [n(%)] | 3(0.69%) | 190(1.26%) | 181(1.99%) | 121(2.39%) | 143(1.94%) | ** |
| Black [n(%)] | 164(38.22%) | 6,128(40.83%) | 3,397(37.49%) | 2,013(39.79%) | 2,893(39.32%) | ** |
| White [n(%)] | 178(41.49%) | 6,680(44.51%) | 3,861(42.61%) | 2,395(47.35%) | 3,702(50.31%) | ** |
| Other Race [n(%)] | 90(20.97%) | 2,008(13.38%) | 1,207(13.32%) | 584(11.54%) | 713(9.69%) | ** |
| **Insurance:** |  |  |  |  |  |  |
| Public [n(%)] | 276(64.33%) | 9,567(63.75%) | 5,407(59.67%) | 2,777(54.90%) | 3,820(51.92%) | ** |
| Private [n(%)] | 153(35.66%) | 4,991(33.26%) | 4,061(44.82%) | 2,270(44.87%) | 3,524(47.89%) | ** |

P-values<sup>a</sup> determined using T-test (continuous variables): \*\* $p < 0.05$ , \* $p < 0.1$ for difference between trajectory clusters;

### Association between the groups and SDOH

We compare the mean values of the SDOH variables using the population mean for each group, comparing it on continuous heatmap gradient, and ANNOVA test across all groups. Figure 3a and 3b show the comparison in a heatmap format for negative and positive SDOHs, respectively. The red color shows group mean is higher and the blue color shows the group mean is lower than the full-cohort mean. At a high level, one can observe that the red boxes are more concentrated on the left side of the 3a, indicating the higher population means in high-risk groups under negative SDOH. The opposite pattern can be observed in 3b. We present a more detailed description of these results in Supplemental-S6.

**Figure 3:**
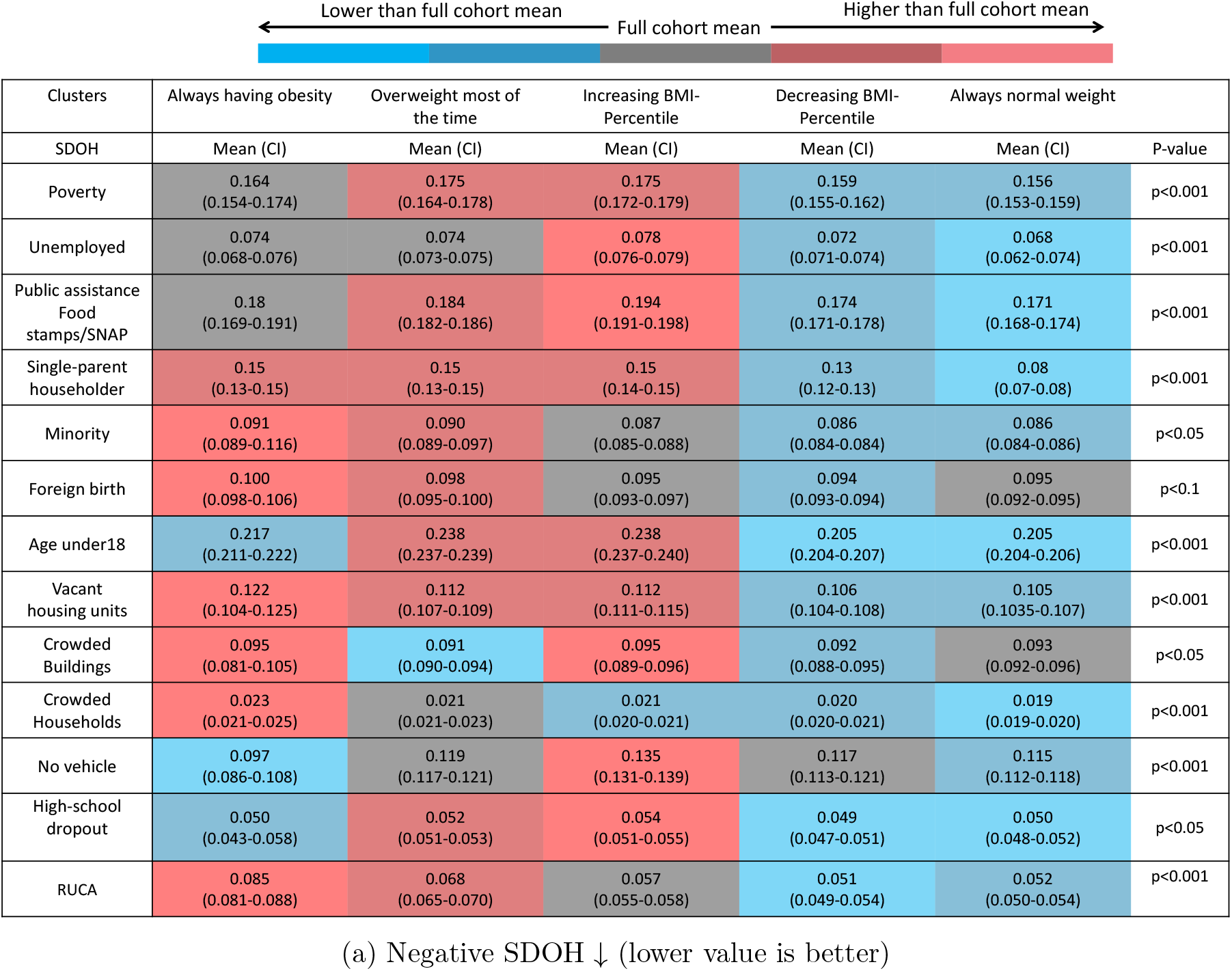
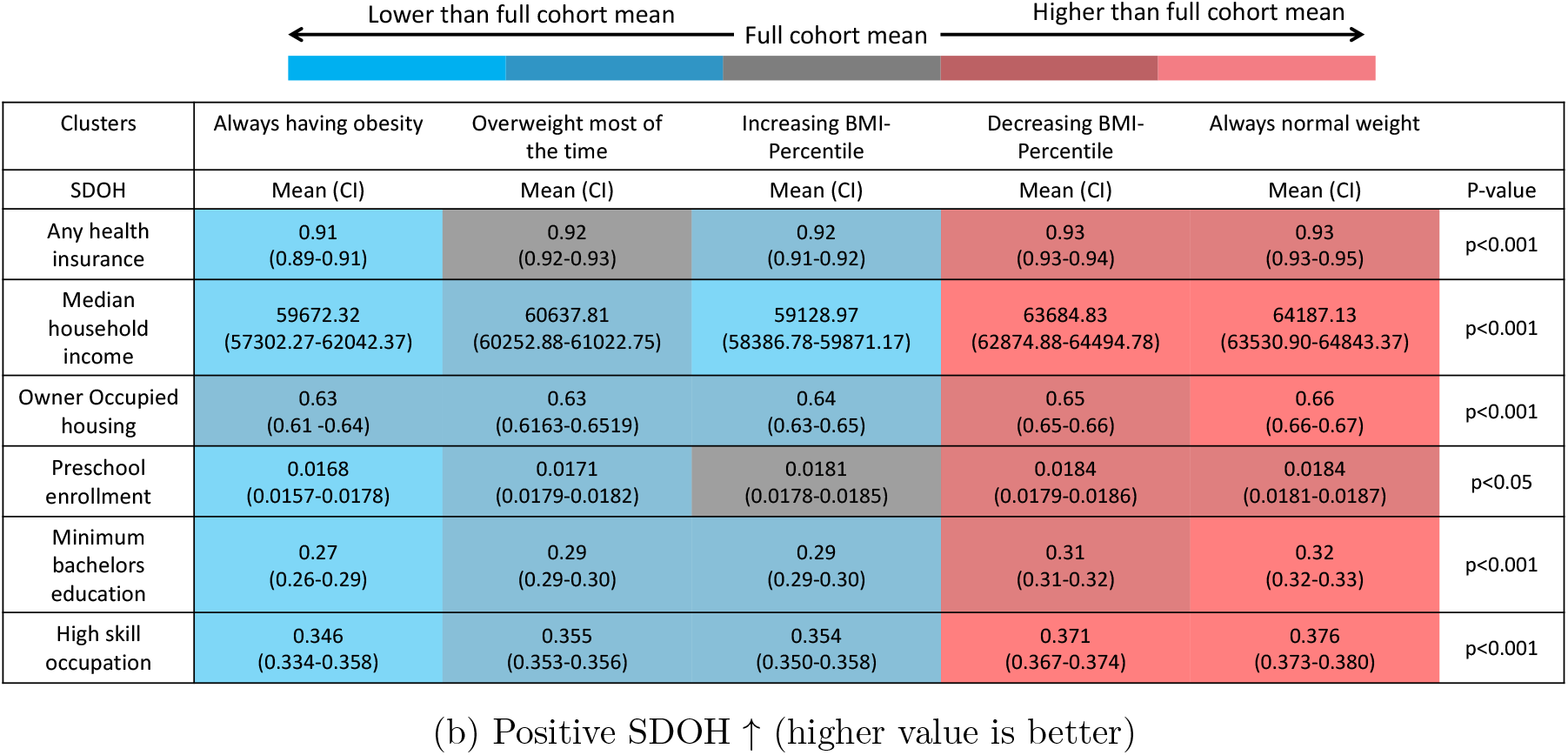
Comparison of mean values for each of the negative (a) and positive (b) SDOH values across the groups using population mean for each group, comparing it on continuous heatmap gradient, and ANNOVA test across all groups. Each column in the table corresponds to a group as labeled in the header. Each row corresponds to the SDOH variable and shows an ANNOVA p-value for that variable across all groups. Each cell shows the population mean value (with 95% CI) for the SDOH variable, and the color shows the visual comparison of the group mean with the total population mean of the SDOH variable. At the top we have a heatmap to help understand the color gradient. The red color shows group mean is higher and the blue color shows group mean is lower compared to the full-cohort population mean. One can especially observe that the higher red heat on the right in (a) is indicative of more negative SDOH factors in the three rightmost high-risk groups and the higher red heat on the left in (b) is indicative of more positive SDOH factors in two low-risk groups identified by our clustering method. The statistical tests were performed using the Researchpy package on Python 3.7.

### Odds of group membership based on the SDOH

Figure 4 shows the results of the multinomial regression for the group membership with ‘normal’ as the reference category. We study the influence of each SDOH listed in Figure 4 on the group membership. The obtained odds ratios show a higher chance of belonging to the higher-risk groups for children with negative SDOH (and vice versa for the positive SDOH). A detailed description of these results is presented in Supplemental-S7.

**Figure 4:**
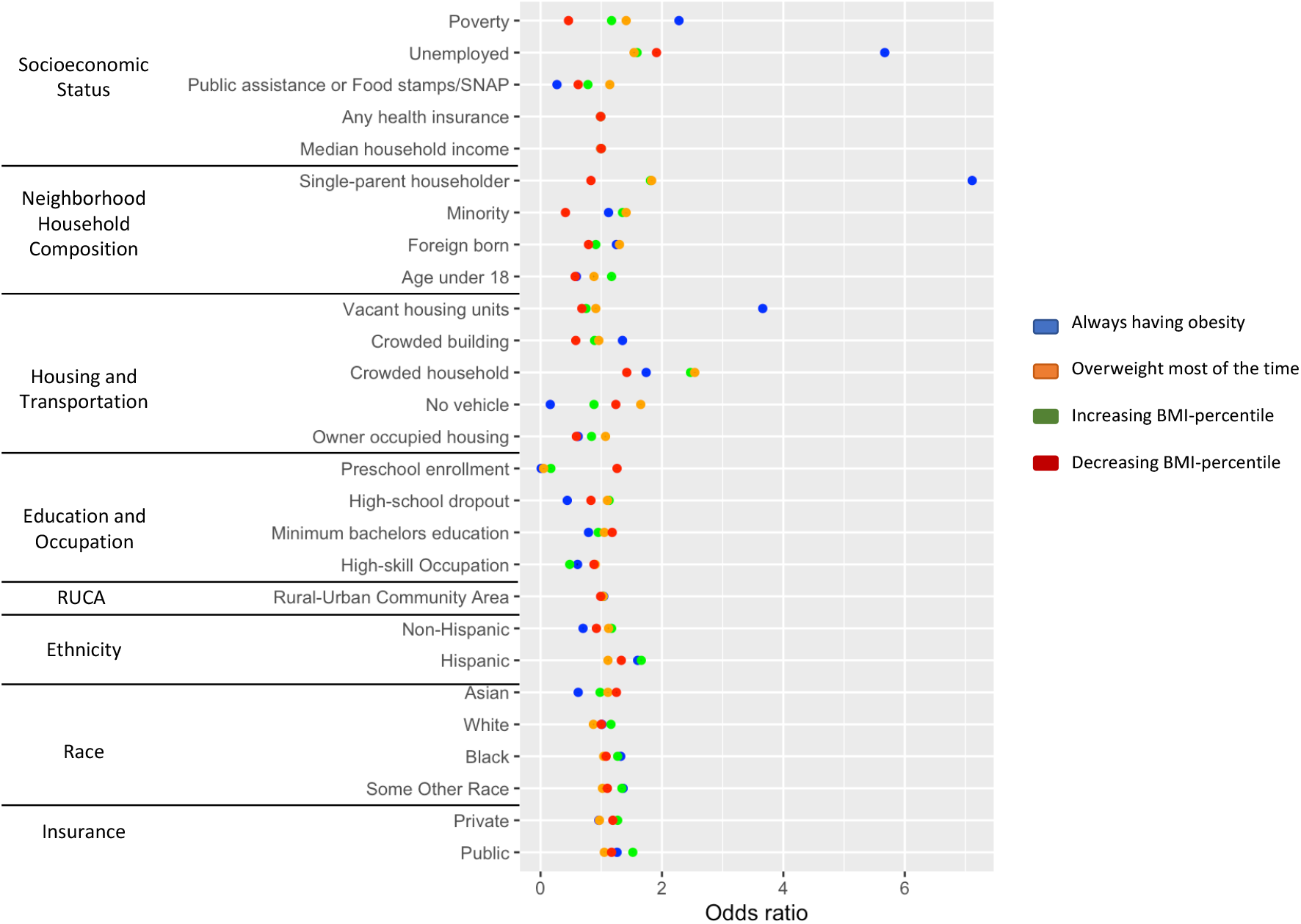
Results from the multivariate logistic regression analysis examining the association of 27 SDOH variables (divided into 8 groups) with the 5 clusters found using our clustering methodology. The units presented in the table are odds ratios. All SDOHs are the ratio of the population belonging to that SDOH except for median household income. RUCA is the ratio of the population with RUCA*≥*4. Additional details about SDOH are shared in Supplemental-S3. Odds ratios using ‘normal (always normal weight)’ as the reference category are shown. Each categorical variable - Non-Hispanic, Hispanic, Asian, White, Black, Some Other Race, Private Insurance, and Public Insurance dummy coded to 0/1 and used the default reference category of 0 for each categorical variable. All CIs did not cross 1 and therefore were significant. Because the CIs were so small, they are not visible on the below plot. All categorical variables in the Ethnicity, Race, and Insurance SDOH groups were dummy-coded to 0/1 and used the default reference category of 0. The analysis was performed using the nnet package from R.

## 4. Discussion

We identified five distinct groups of BMI-percentile classification trajectories among 36,910 children from birth to 7 years of age. Two groups demonstrated stable BMI-percentile classifications over time, one with a BMI-percentile always above 95% (‘obesity’) and another with a BMI-percentile always less than 85% (‘normal’). Using algorithmic clustering, we also found three additional groups - two groups showed an increasing trajectory, with one group (‘overweight’) showing a smaller and the other (‘increasing’) showing a larger increase in BMI-percentile. A third group (‘decreasing’) started with a BMI-percentile near 85% but then demonstrated a downward trajectory into a normal BMI-percentile range (*<*85%).

These five groups are consistent with the groups found in other longitudinal studies that find different BMI trajectories in children.^37,40,51,68–70^ However, the majority of these studies look at trajectories, without a relationship to widely-established BMI-percentile classifications. Our identification of a similar five groups, but in relationship to well-established and clinically-used BMI-percentile classifications will allow clinicians to better interpret the findings of our study. As an example, it would be easier for a clinician to interpret the higher social risks for children if they are categorized as not only having an increasing weight but going from normal weight to overweight or obesity.

Some of these trajectory studies have demonstrated associations between increasing trajectories and individual-level environmental factors like food and housing insecurity and maternal social risk factors like depression and substance use.^51,71–73^ Some other studies have examined the association between neighborhood-level SDOH and risk for overweight and obesity,^74–77^ but only using cross-sectional data.^13,45,78–81^ Our study is distinct in that it includes the analysis of the association between 27 different social and neighborhood-level SDOH risk factors and five distinct BMI-percentile trajectories based on changes in clinically-relevant BMI-percentile classifications during early childhood.

We find three high-risk groups (‘obesity’, ‘overweight’, and ‘increasing’) with distinct BMI-percentile trajectories and SDOH. This is an important finding of our study because these differences among high-risk groups can be used to inform tailored interventions for each group. Those in the ‘obesity’ and ‘overweight’ groups – children who demonstrated very early onset overweight or obesity from infancy – were more likely to be from neighborhoods with more crowded buildings and households, with a higher percentage of the population from Hispanic and immigrant backgrounds. It, therefore, seems that providing targeted intervention to Hispanic and immigrant communities in historically under-resourced neighborhoods, especially providing support in the perinatal period, supporting pregnant women and their newborns, and addressing both maternal lifestyle and infant feeding practices, may be important to preventing overweight and obesity in infancy. Whereas the ‘increasing’ group, who developed obesity over time, were more likely to be from neighborhoods with a higher Black population, with higher rates of unemployment, high-school dropouts, and dependence on public assistance. Therefore, providing targeted interventions to Black communities in historically under-resourced neighborhoods to reduce inequities in resources such as employment, and education would be critical to preventing rapid weight gain that might occur in early childhood.

Numerous other studies have shown, factors like socioeconomic-status, and household structures can also contribute significantly to obesogenic lifestyles.^82–88^ Beyond that, neighborhoods can also determine access to healthcare, education, and transportation and one’s social environment (e.g., norms, relationships, stressors) can negatively impact an individual’s self-efficacy for engaging in health promotion behaviors.^89–93^ These barriers disproportionately impact communities of color, who are more likely to live in historically under-resourced neighborhoods due to redlining and lack of investment in their communities from decades of systemic racism.^94^ Therefore, for all of these high-risk groups, it would be critically important to develop effective interventions for reducing obesity-related disparities driven by SDOH, and help families overcome barriers to care, such as food insecurity, poverty.^15, 95–97^

Interestingly, our study also found a group with a decreasing BMI-percentile. In direct contrast to increasing BMI-percentile groups, we found that children with decreasing trajectories were more likely to be from neighborhoods with higher rates of preschool enrollment and bachelor’s education, and lower rates of poverty, reliance on public assistance, single-parent households, and crowded households. This is consistent with other studies that demonstrate the importance of education and SES to weight loss.^98–100^ While we did not intend to look at children who can successfully lose weight with this study, our ability to distinguish this group with clustering and our finding that there were SDOH associations in the opposite direction as our groups with obesity are very telling about the important role of SDOH not only to the development of obesity but how successful families might be if provided with more social and environmental resources.

## 5. Limitations

Our study is limited in several ways. For linking patients to one census-tract, we have used their last available address during the 0–7 years. While relocations were not frequent, a more advanced linkage by tracking all the locations over time could enhance this process. We excluded children with very few observations to ensure our analysis of trajectories was reliable, there may be selection bias since children from households with lower socioeconomic-status may be more likely to move out of the healthcare system. However, 42% of patients had public insurance in our final dataset and we were still able to rigorously evaluate the contribution of socioeconomic factors on weight trajectories with our dataset. Our study cohort is very large, spans several US states, and is collected across all settings within the healthcare system and specialties. While this increases the generalizability of the results, there may be regional differences that we did not account for. In addition, there may be some limitations to not excluding certain populations like those on certain medications that impact weight, those with monogenic obesity, or those seeking weight management treatment. However, for our primary aim of evaluating SDOH on weight trajectories, we felt it was important to be inclusive of most of the population to best understand this question but in future studies would consider a stratified analysis based on these factors. Finally, this analysis did not consider the potential interactions between the different SDOH variables studied. Future studies should employ approaches like multilevel analysis of individual heterogeneity and discriminatory accuracy to better understand these intersectionalities.

## 6. Conclusion

Our findings, leveraging a large EHR dataset linked to neighborhood-level SDOHs and clustering, found five groups that differed in how their BMI-percentile classification changed throughout early childhood. Three groups demonstrated high-risk of developing obesity and were more likely to be from historically under-resourced neighborhoods. Importantly, these three groups were different from one another in terms of the timing of the development of overweight and obesity which supports the need to develop tailored interventions for both perinatal and early childhood obesity prevention.

## Data Availability

All the pre-processed data produced in the present study can be made available upon reasonable request to the authors and upon signing a data use agreement.

## Abbreviations

BMI: Body Mass Index
BMI-percentile: Body Mass Index percentile
EHR: Electronic Health Record
SDOH: Social Determinants of Health

## Authors’ Contributions

MG conceived and carried out experiments. TP and RB conceived the experiments and helped in the study design. MG, FL, DE, and HB worked on preprocessing steps including data extraction, cleaning, and linkage. All authors were involved in writing the paper and had final approval of the submitted and published versions.

