## supplementary material for "Associations of longitudinal BMI percentile classification patterns in early childhood with neighborhood-level social determinants of health"

### Supporting information

#### Section S1:

#### Using BMI percentile in all ages

We note that the American Academy of Pediatrics recommends weight-for-length (WFL) for children younger than 2 yrs, but many recent studies show that BMI has a higher predictive value for childhood obesity and other obesity-related comorbidities than WFL.[1-3] Both BMI-percengtile and WFL-percentile use anthropometric measurements of height and weight and demographic variables of age and sex. To remain consistent with the measurements used for the cohort across all time periods (0-7 years), and consistent with many other group-based trajectory studies that have used BMI from birth to ages above 2 years to find the growth trajectories, we chose to use BMI% across all ages[1,4-6].

#### Section S2:

We calculated the number of children in different BMI- percentile trajectories as shown in Table S1. The tables shows that there were no children with BMI- percentile always below 5. Also, there were no child who had both BMI- percentile below 5 and BMI- percentile above 85 (row 9) or above 95 (row 8). However there were children whose BMI- percentile fluctuates between below 5 and between the 5 to 85 thresholds. Because of the focus of our study on children with overweight or obesity (i.e., above 85 and 95 respectively), we excluded the children in row 10. There were only 7 children with BMI- percentile always between 85 to 95, we excluded the children in row 2, too.
Childern in row 5 and row 6 overlap with children in row 7 which is included in our study. Children in row 1 and row 4 are also included in the study.

**Table S1:** The number of children in different BMI- percentile thresholds. Blue rows refer to those we included in our study.

| | **Number of children with BMI-** percentile **trajectories** | **Count** |
|---|---|---|
| 1 | Always above 95% | 429 |
| 2 | Always between 85 to 95 (overweight BMI%) | 7 |
| 3 | Always below 5 (underweight BMI%) | 0 |

| | | |
|---|---|---|
| 4 | Always between 5 to 85 (normal BMI%) | 7357 |
| 5 | BMI% recorded from both overweight BMI% (between 85 to 95) and obesity BMI% (above 95) | 1569 |
| 6 | BMI% recorded from both normal BMI% (between 5 to 85) and obesity BMI% (above 95) | 1169 |
| 7 | BMI% recorded from at least two of the normal BMI% (between 5 to 85), overweight BMI% (between 85 to 95) and obesity BMI% (above 95) | 29124 |
| 8 | BMI% recorded from both underweight BMI% (below 5) and obesity BMI% (above 95) | 0 |
| 9 | BMI% recorded from both underweight BMI% (below 5) and overweight BMI% (between 85 to 95) | 0 |
| 10 | BMI% recorded from both underweight BMI% (below 5) and normal BMI% (between 5 to 85) | 3976 |

### Section S3:

#### Geolocating SDOH Data

The data consisted of clinical information and demographic information such as race (categorized for the purpose of analysis as White, Black, Asian, and Other) and ethnicity (categorized for the purpose of analysis as Hispanic and Non-Hispanic), and insurance type (categorized for the purpose of analysis as private and public).

Patient addresses were mapped to latitude, longitude, and census tract codes using the Census Geocoder. The Census Geocoder address look-up tool allowed us to map 3,012,495 of 3,488,128 patient visits (86.36%) to 4,237 unique census tract geocodes. On average, each patient was mapped to 1.27 different census tracts from the first visit to the last visit of this study. 23% of patients had a relocation, out of which 58% had no relocation after 3 years of age showing that their latest census-tract is also the longest-stayed census-tract. We used the latest census-tract for each patient.

For linking the above cohort to SDOH data, we initially compiled a repository of socioeconomic data from the 2005 to 2018 U.S. Census Bureau American Community Survey (ACS) 5-year estimates[7] and from the 2010 U.S. Department of Agriculture Rural-Urban Commuting Area (RUCA) codes.[8] The 2005 to 2009 ACS survey data was mapped to 2000 census tract boundaries with 66,332 geographic IDs and 868 socio-economic and demographic fields for each census tract. The 2010 to 2018 ACS survey data and 2010 RUCA codes were mapped to 2010 census tract boundaries with 74,001 geographic IDs with 951 socio-economic and demographic fields for each census tract. The 13.64% of missing codes were the result of data entry errors, or no longer existing addresses.

To preserve patient confidentiality and prevent exposure of patient-protected health information (PHI), census tracts were mapped to randomly generated codes that allowed the linking of patients to socio-economic data by a designated honest broker.

We identified the unique variables from the Cesnus American Community Survey (ACS) that are used in these indices and grouped them into five categories: 1) socioeconomic status, 2) neighborhood household composition, 3) housing and transportation, 4) education and occupation, and 5) RUCA code. The socioeconomic status category included the following variables: poverty - % of persons below the poverty level, unemployed - % of unemployed persons over age 18 in the neighborhood, public assistance or food stamps/SNAP - % households receiving public assistance, any health insurance – % persons with any health insurance (all ages), and median household income. The neighborhood household composition category included single householder - % single caregiver household, % minority (race or ethnicity other than Non-Hispanic White), % foreign-born, and age under 18 - % age less than 18. The housing and transportation category included vacant housing units - % vacant housing units, crowded buildings - % crowded buildings (buildings with >= 10 units), crowded households - % crowded households (occupied housing units with > 1 person per room), no vehicle - % of housing units with no vehicle available, and owner-occupied housing - % housing units owner-occupied, among the occupied housing units. The education and occupation category included preschool enrollment - % children age 3-6 in nursery school or preschool, high-school dropout - % of persons 16-19 years of age not enrolled in or graduated from high school, minimum bachelors education - % of persons age 25 to 34 years of age with a bachelor's degree), high skill occupation - % of persons above 25 years of age with a high skill occupation (management, business, science, and arts). All these variables are represented as ratios instead of percentages in our experiments. Finally, the RUCA category includes RUCA codes, which describe the degree of urbanization of a neighborhood based on characteristics like population density and distance between households on a scale of 1 to 10 with neighborhoods having a RUCA code of >= 4 being more rural. We represented RUCA also as the ratio of the population in cohort with population ratio belonging to RUCA>=4 in each group and cohort.

### Section 4:

### Clustering Approach

We used a two-step process to identify BMI% trajectory groups. We first used a data-driven approach where we used BMI% data points in each patient's history to find any homogeneous groups who remained in the same BMI% category over time. We found two such groups: 1) BMI% always in normal weight range (5<= BMI% < 85), 2) BMI% always in the obesity range (BMI% >= 95). The remaining group thus represented a group with significant heterogeneity, with individuals in that group having a BMI% recorded from at least two of the normal, overweight, or obesity categories (5<= BMI%). We used partitional clustering with DTW distance to identify clusters in all these three groups.

Partitional clustering is a stochastic algorithm that partitions data into a certain number of clusters, with a centroid for each cluster. Using this method, first, k cluster centroids are randomly initialized with k randomly selected time series in the data. The distance between each series and each centroid is calculated. Here, we use dynamic time warping (DTW) to calculate the distance between time series. The time series is assigned to the cluster whose centroid is closest. This procedure is repeated until no time series changes from one cluster to another.

DTW is a method of aligning two time-series sequences to calculate distance based on their pattern similarity.[9] Before applying DTW methodology to calculate distance, we interpolated BMI% time series to obtain continuous signals of 7 years with data distributed over 3-month time periods. We used the *imputeTS R package*,[10] which specializes in univariate time-series imputation, with the linear interpolation setting to impute the missing BMI% values for children (results shown in Figure S1). With a continuous time-series distributed over 3-month intervals and the warping window restrictions of the warping function of DTW the alignment calculated between two time-series using DTW does not wander too far from the diagonal. The DTW distance between each trajectory was used to find the groups of trajectories using partitional clustering. We identified a representative trajectory curve for each group as the centroid of each group. We used Bayesian Information Criteria and sample size to be at least 10% of the input sample size as our criteria to select the number of clusters. Using this cirteria, we only find clusters in third group. Figure S2 shows BIC with number of clusters for all groups. We identify 3 clusters in third group. We combined three clusters from third group and other two homogeneous groups. Figure S3 shows all the 5 clusters. 3 clusters identified using clustering approach are – overweight, increasing and decreasing.

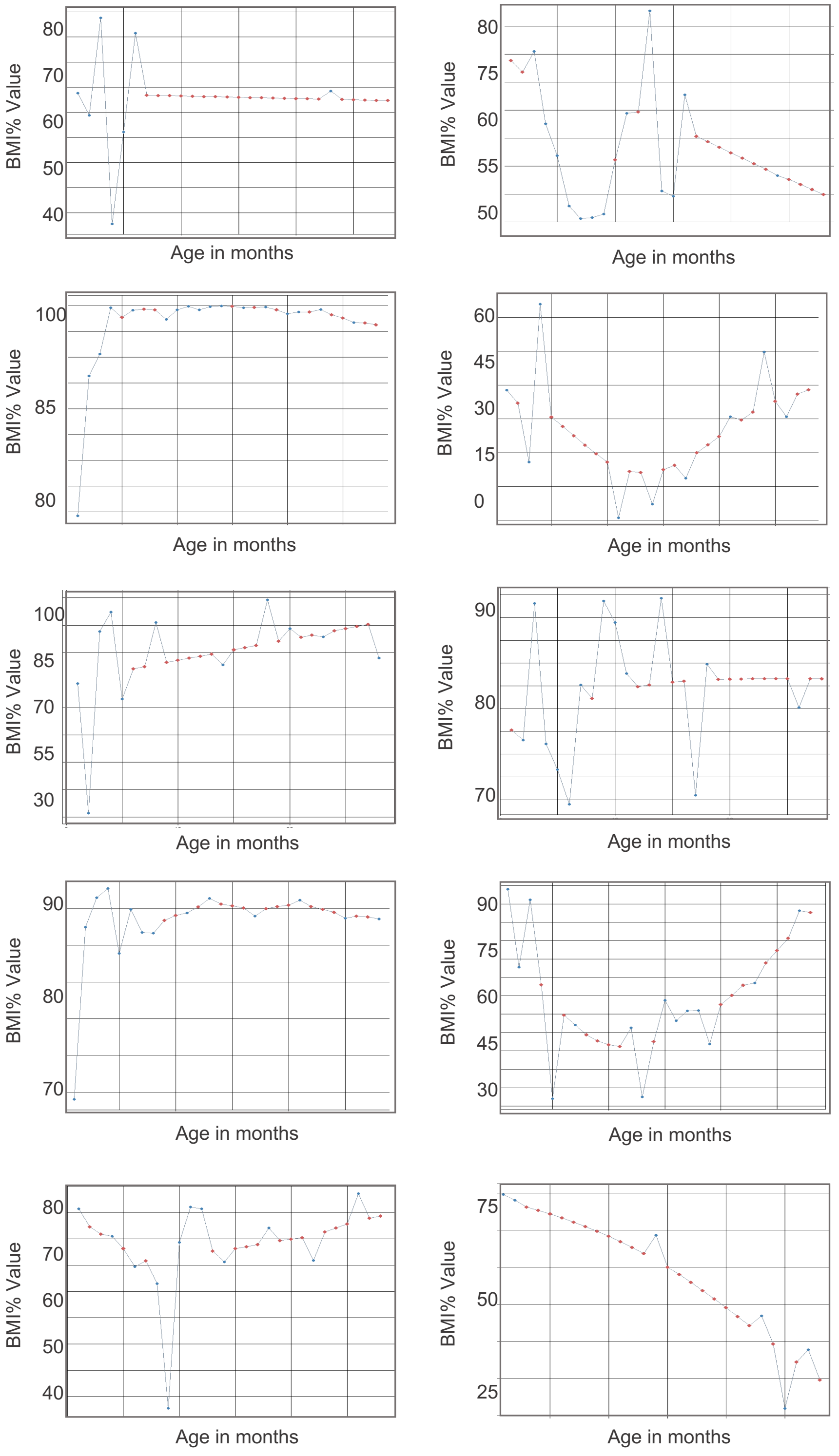


**Figure S1**: Randomly selected BMI% trajectories showing the recorded values (blue) and the imputed values (red) obtained from time-series imputation to create continous BMI% trajectories.

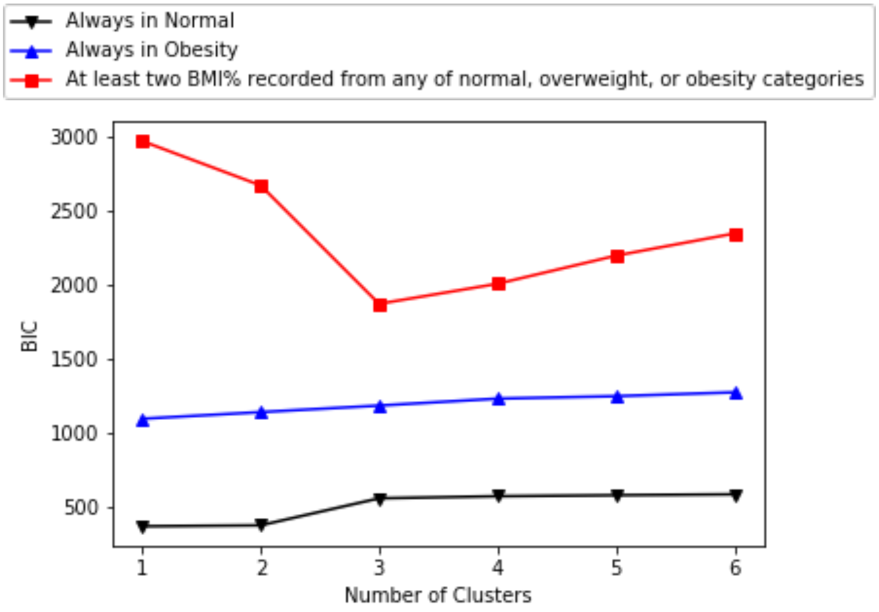


**Figure S2:** 1) BMI% always in normal weight range, 2) BMI% always in the obesity range, and 3) at least two BMI% recorded from any of normal, overweight, or obesity categories. BIC: Bayesian Information Criteria.

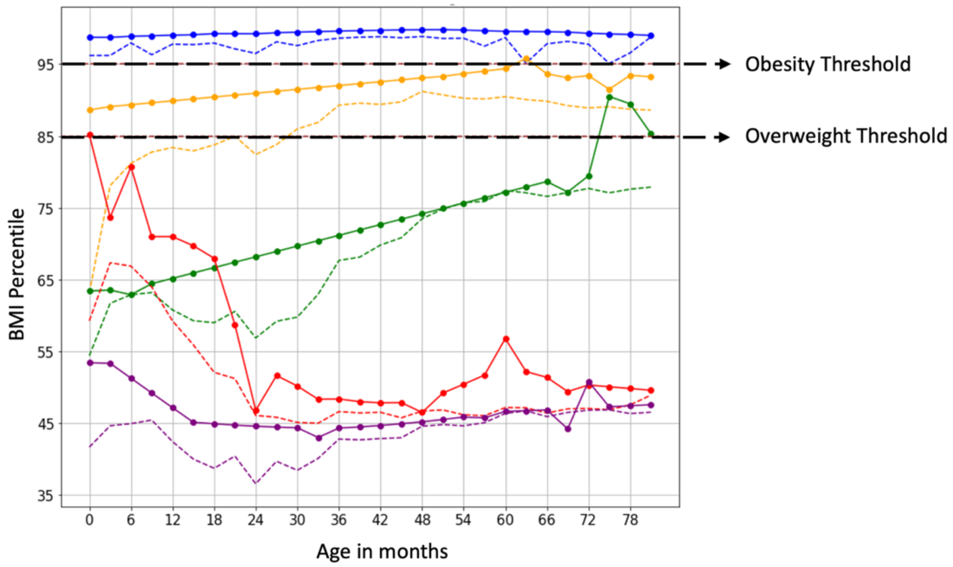


**Figure S3:** Five groups were identified in our study based on BMI% trajectories of children between 0 to 7 years. **Solid lines** show the representative (centroid - center of the cluster) BMI% trajectories for each group. **Dotted lines** show the mean BMI% trajectories for each group.

### Section S5:

### Applying LCGMM for clustering heterogeneous group of BMI% trajectories

We also verified our results from the partitional clustering approach using an alternative, group-based trajectory method. LCGMM is a longitudinal technique that can be used to capture the heterogeneity in BMI% trajectories over time. We compared linear and quadratic LCGMM techniques to choose a final model. To determine the optimal number of classes, we compare models by starting from one class and adding more classes one at a time. We use Bayesian Information Criterion (BIC), Bootstrapped Likelihood Ratio Test (BLRT), and posterior probabilities to compare the fit of different models.[11,12] The BIC considers both likelihood of the model as well as the number of parameters in the model.[13] The BLRT provides a p-value, comparing a model with k-1 classes with a model with k classes.[14,15] The posterior probability is used to assign children to their most-likely class. A higher difference between the probability of an assigned class and the probability of a class not assigned makes the classes more distinguishable. Table S2 shows the results from the experiments we ran to determine the number of classes.

**Table S2:** Model fit indices of LCGMM

| Number of Classes | Bayesian Information Criterion | Average Posterior Probability | P-value | Number of subjects per class |
|---|---|---|---|---|
| 1 | 319942.56 | 1.00 | - | |
| 2 | 319915.32 | 0.753 | $p<0.001$ | 20,234/8,890 |
| 3 | 319898.59 | 0.832 | $p<0.001$ | 15,006/9,060/5,058 |
| 4 | 319908.73 | 0.772 | $p<0.001$ | 16,133/6,892/2,523/3,576 |

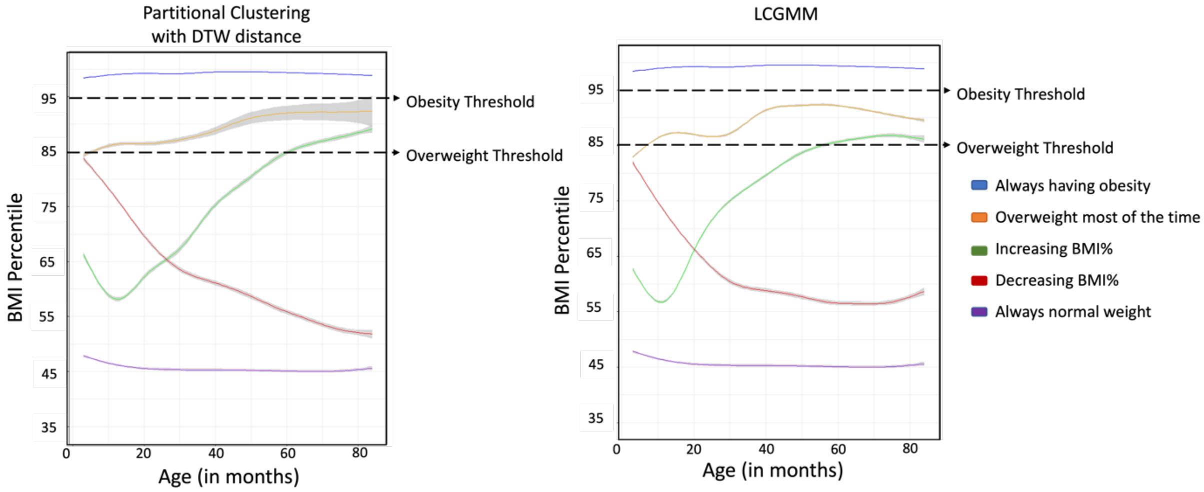


**Figure S4:** Comparison of five groups identified in our study based on BMI% trajectories of children between 0 to 7 years. Left are the clusters identified by partitional clustering using DTW distance. Right are the clusters identified by LCGMM. We used a smoothing function (geom_smooth package in R) to represent each cluster by a smooth solid line with 95% CI.

### Section S6:

### Association between the clusters and SDOH

As Figure 3A in the main manuscript show, children from the clusters indicating higher concerns for obesity (`obesity', `overweight', and `increasing') have more concentration of red in negative SDOH areas. Red color shows that mean of population in these groups is higher compared to full-cohort mean and mean of low risk groups. High-risk groups were from neighborhoods with a higher number crowded households ($p<0.001$) and individuals of lower socioeconomic status ($p<0.001$), higher minority population ($p<0.05$), single-parent households ($p<0.001$), and greater rurality ($p<0.001$).

An opposite trend is observed in Figure 3B which shows comparison for positive SDOH across groups. Higher concentration of blue in high-risk groups in Figure 3B shows that mean of population in these groups is lower in positive SDOH areas compared to full-cohort mean and mean of low-risk groups. High-risk groups have lower educational status with high high-school dropout ($p<0.05$) and low preschool enrollment ($p<0.05$).

Furthermore, children from the groups indicating lower concerns for obesity (`normal', and `decreasing') have higher socioeconomic status with lower poverty ($p<0.001$) and unemployment ($p<0.001$), higher owner-occupied housing ($p<0.001$), higher education with higher preschool enrollment ($p<0.05$), and bachelor's educated ($p<0.001$).

### Section S7:

### Odds of cluster membership based on the SDOH

We have added Table S3 in this section which shows detailed odds-ratio for the plot shown in Figure 4 in the main manuscript. For Table S3 and Figure 4 we can see that children from the higher SDOH risk factors had greater odds of being in `obesity' group compared to the reference category (`normal'), but the odds are especially higher for neighborhoods with more households with incomes below the poverty level (Odds Ratio 2.3), more unemployment (OR 5.7), more single-parent households (OR 7.1), more vacant housing units (OR 3.6), crowded buildings (OR 1.3), and rurality (OR 1.04). The odds of being in the `overweight' group are greater for neighborhoods with higher high-school dropouts (OR 1.13), with more persons from racial and ethnic minority backgrounds (OR 1.3), particularly Hispanic ethnicity (OR 1.6), and having public insurance (1.5). Finally, the odds of being in the `increasing' group are higher for neighborhoods with crowded households (OR 2.5), more population from minority (OR 1.41), foreign-born (OR 1.30), and those who rely on public assistance (OR 1.14). However, the odds are lower for `obesity', and `overweight' groups with increasing preschool enrollment (OR 0.006, 0.1675), bachelor's education (OR 0.79, 0.95), and high-skill occupation (OR 0.61, 0.48), respectively. In contrast, we observe lower odds of being in the `decreasing' group compared to the baseline group `normal' with increasing poverty (OR 0.46), public assistance (OR 0.62), single householders (OR 0.83), crowded buildings (OR 0.58), and high school dropout (OR 0.83) and the odds increase with increasing preschool enrollment (OR 1.26) and bachelor's education (OR 1.18).

**Table S3:** Results from the multivariate logistic regression analysis examining the association of 27 SDOH variables (divided into 8 groups) with the 5 clusters found using our clustering methodology. The units presented in the table are odds ratios. All SDOHs are the ratio of the population belonging to that SDOH except for median household income. RUCA is the ratio of the population with RUCA>=4. Odds ratios using `normal (always normal weight)' as the reference category are shown. Each categorical variable - Non-Hispanic, Hispanic, Asian, White, Black, Some Other Race, Private Insurance, and Public Insurance are dummy coded to 0/1 and used the default reference category of 0 for each categorical variable. All CIs did not cross 1 and therefore were significant. All categorical variables in the Ethnicity, Race, and Insurance SDOH groups were dummy coded to 0/1 and use the default reference category of 0.

| SDOH group | SDOH | Obesity (Always having obesity) | Overweight (Overweight most of the time) | Increasing (Increasing BMI-percentile) | Decreasing (Decreasing BMI-percentile) |
|---|---|---|---|---|---|
| Socioeconomic Status | Poverty | 2.28 | 1.17 | 1.41 | 0.46 |
| | Unemployed | 5.67 | 1.59 | 1.54 | 1.91 |
| | Public assistance or Food stamps/SNAP | 0.27 | 0.78 | 1.14 | 0.62 |
| | Any health insurance | 0.99 | 0.99 | 0.99 | 0.99 |
| | Median household income | 0.99 | 1.00 | 0.99 | 1.00 |
| Neighbourhood Household Composition | Single-parent householder | 7.11 | 1.81 | 1.83 | 0.83 |
| | Minority | 1.12 | 1.35 | 1.41 | 0.41 |
| | Foreign born | 1.25 | 0.91 | 1.30 | 0.79 |
| | Age under 18 | 0.59 | 1.17 | 0.88 | 0.57 |

| | | | | | |
|---|---|---|---|---|---|
| Housing and Transportation | Vacant housing units | 3.66 | 0.75 | 0.91 | 0.68 |
| | Crowded building | 1.35 | 0.89 | 0.96 | 0.58 |
| | Crowded household | 1.74 | 2.47 | 2.54 | 1.42 |
| | No vehicle | 0.16 | 0.88 | 1.65 | 1.24 |
| | Owner occupied housing | 0.62 | 0.84 | 1.07 | 0.59 |
| Education and Occupation | Preschool enrollment | 0.0006 | 0.17 | 0.05 | 1.26 |
| | High-school dropout | 0.44 | 1.13 | 1.10 | 0.83 |
| | Minimum bachelor's education | 0.79 | 0.95 | 1.05 | 1.18 |
| | High-skill Occupation | 0.61 | 0.48 | 0.90 | 0.88 |
| Rural-Urban | Rural-Urban Community Area | 1.04 | 1.03 | 1.03 | 0.99 |
| Ethnicity | Non-Hispanic | 0.70 | 1.17 | 1.12 | 0.92 |
| | Hispanic | 1.60 | 1.66 | 1.11 | 1.33 |
| Race | Asian | 0.62 | 0.98 | 1.11 | 1.25 |
| | White | 1.01 | 1.16 | 0.87 | 1.00 |
| | Black | 1.32 | 1.27 | 1.04 | 1.08 |
| | Some Other Race | 1.36 | 1.34 | 1.02 | 1.10 |
| Insurance | Private | 0.96 | 1.27 | 0.97 | 1.19 |
| | Public | 1.26 | 1.52 | 1.05 | 1.17 |

**Section S8:**

**Disease included under Cancer:**

'Malignant tumor of submandibular gland', 'Malignant tumor of oral cavity', 'Primary malignant neoplasm of larynx', 'Primary malignant neoplasm of pineal gland', 'Secondary malignant neoplasm of pleura', 'Primary malignant neoplasm of pelvic bones, sacrum and coccyx', 'Malignant neoplasm of lateral wall of urinary bladder', 'Malignant neoplasm of posterior wall of urinary bladder', 'Primary malignant neoplasm of bone', 'Malignant neoplasm of anterior wall of urinary bladder', 'Secondary malignant neoplasm of bone', 'Secondary malignant neoplasm of urinary system', 'Primary malignant neoplasm of scrotum', 'Primary malignant neoplasm of anus', 'Malignant neoplasm of thorax', 'Chronic myeloid leukemia in remission', 'Myeloid sarcoma in remission', 'Malignant lymphoma of lymph nodes of multiple sites', 'Myeloid leukemia in remission', 'Lymphoid leukemia', 'Primary malignant neoplasm of skin of trunk', 'Plasma cell leukemia', 'Plasma cell leukemia in remission', 'Primary malignant neoplasm of endocrine gland', 'Primary malignant neoplasm of thyroid gland', 'Chronic lymphoid leukemia in remission', 'Lethal midline granuloma', 'Overlapping malignant neoplasm of female breast', 'Malignant melanoma of skin of face', 'Malignant melanoma of skin of trunk', 'Acute lymphoid leukemia', 'Lymphoid leukemia in remission', 'Chronic leukemia in remission', 'Chronic myeloid leukemia', 'Lymphosarcoma of intrathoracic lymph nodes', 'Mycosis fungoides of extranodal AND/OR solid organ site', 'Polycythemia vera', 'Primary malignant neoplasm of spinal cord', 'Acute leukemia in remission', 'Subacute myeloid leukemia', "Sézary's disease of extranodal AND/OR solid organ site", 'Acute myeloid leukemia in remission', 'Mycosis fungoides of lymph nodes of multiple sites', 'Leukemia in remission', 'Acute monocytic leukemia', 'Chronic monocytic leukemia', 'Secondary malignant neoplasm of skin', "Hodgkin's disease, mixed cellularity of intrathoracic lymph nodes", 'Subacute lymphoid leukemia', "Hodgkin's disease, nodular sclerosis of intrathoracic lymph nodes", 'Megakaryocytic leukemia in remission', 'Myelodysplastic syndrome with isolated del(5q)', 'Erythroleukemia, FAB M6', 'Mycosis fungoides of lymph nodes of head, face AND/OR neck', 'Chronic lymphoid leukemia, disease', 'Acute leukemia', 'Myelodysplastic syndrome', 'Primary malignant neoplasm of skin', 'Malignant melanoma of skin of lower limb', 'Chronic leukemia', 'Myelosclerosis with myeloid metaplasia', 'Acute myeloid leukemia, disease', "Hodgkin's paragranuloma of lymph nodes of multiple sites", 'Myeloid leukemia', 'Acute monocytic leukemia in remission', 'Myeloid sarcoma', 'Malignant melanoma of skin', "Hodgkin's disease, lymphocytic depletion of intrathoracic lymph nodes", 'Letterer-Siwe disease of lymph nodes of multiple sites', 'Acute lymphoid leukemia in remission', "Hodgkin's disease, mixed cellularity of intra-abdominal lymph nodes", 'Malignant lymphoma of lymph nodes of inguinal region AND/OR lower limb', 'Secondary malignant neoplasm of intra-abdominal lymph nodes', 'Secondary malignant neoplasm of adrenal gland', "Hodgkin's disease, nodular sclerosis of spleen", "Burkitt's lymphoma of intrapelvic lymph nodes", "Hodgkin's disease, nodular sclerosis of intra-abdominal lymph nodes", 'Malignant neoplasm of corpus uteri, excluding isthmus', 'Secondary malignant neoplasm of inguinal lymph nodes', "Hodgkin's disease, lymphocytic-histiocytic predominance of intrapelvic lymph nodes", 'Nodular lymphoma of extranodal AND/OR solid organ site', 'Malignant lymphoma of intrapelvic lymph nodes', "Hodgkin's disease, mixed cellularity of lymph nodes of inguinal region and lower limb", "Hodgkin's disease of intrapelvic lymph nodes", 'Secondary malignant neoplasm of kidney', "Burkitt's lymphoma of lymph nodes of inguinal region and lower limb", 'Primary malignant neoplasm of bladder', 'Malignant tumor of kidney', 'Secondary malignant neoplasm of retroperitoneum and peritoneum', 'Malignant neoplasm of uterus', 'Primary malignant neoplasm of colon', 'Malignant neoplasm of abdomen', 'Primary malignant neoplasm of male genital organ', 'Malignant tumor of urinary bladder', 'Primary malignant neoplasm of gallbladder', 'Malignant neoplasm of uterine adnexa', 'Letterer-Siwe disease of spleen', "Kaposi's sarcoma of gastrointestinal tract", 'Primary malignant neoplasm of retroperitoneum', 'Overlapping malignant neoplasm of retroperitoneum and peritoneum', 'Secondary malignant neoplasm of small intestine', 'Secondary malignant neoplasm of liver', 'Lymphosarcoma of lymph nodes of inguinal region and lower limb', "Hodgkin's disease of spleen", 'Malignant tumor of cervix', 'Primary malignant neoplasm of kidney', 'Primary malignant neoplasm of pelvis', "Hodgkin's disease, nodular sclerosis of intrapelvic lymph nodes", 'Carcinoid syndrome', 'Primary malignant neoplasm of uterine adnexa', 'Nodular lymphoma of intra-abdominal lymph nodes', 'Malignant lymphoma of spleen', 'Secondary malignant neoplasm of large intestine', 'Malignant lymphoma of intra-abdominal lymph nodes', "Hodgkin's disease of intra-abdominal lymph nodes", 'Malignant mast cell tumor of intrapelvic lymph nodes', 'Secondary malignant neoplasm of intrapelvic lymph nodes', 'Primary malignant neoplasm of prostate', 'Malignant neoplasm of connective and soft tissue of pelvis', "Hodgkin's disease, lymphocytic-histiocytic predominance of intra-abdominal lymph nodes", 'Primary malignant neoplasm of lower limb', 'Primary malignant neoplasm of liver', 'Primary malignant neoplasm of upper respiratory tract', 'Secondary malignant neoplasm of respiratory tract', "Kaposi's sarcoma of lung", 'Secondary malignant neoplasm of lung', 'Malignant neoplasm of bones of skull and face', 'Primary malignant neoplasm of lung', 'Overlapping malignant neoplasm of bronchus and lung', "Kaposi's sarcoma of skin", 'Primary malignant neoplasm of accessory sinus', 'Megakaryocytic leukemia', "Hodgkin's disease, lymphocytic depletion of lymph nodes of multiple sites", "Hodgkin's disease, nodular sclerosis of lymph nodes of axilla AND/OR upper limb", "Hodgkin's disease, mixed cellularity of lymph nodes of multiple sites", "Burkitt's lymphoma of lymph nodes of multiple sites", "Hodgkin's disease, nodular sclerosis of lymph nodes of multiple sites", "Hodgkin's disease of lymph nodes of multiple sites", 'Primary malignant neoplasm of heart', 'Leukemia', 'Primary malignant neoplasm of soft tissues of thorax', 'Secondary malignant neoplasm of lymph node', 'Lymphosarcoma of lymph nodes of multiple sites', "Hodgkin's granuloma of lymph nodes of multiple sites", 'Secondary malignant neoplasm of lymph nodes of multiple sites', 'Nodular lymphoma of lymph nodes of multiple sites', "Hodgkin's disease, lymphocytic-histiocytic predominance of lymph nodes of multiple sites", 'Monocytic leukemia', 'Malignant histiocytosis of lymph nodes of multiple sites', 'Malignant neoplasm of nasolacrimal duct', 'Primary malignant neoplasm of conjunctiva', 'Primary central nervous system lymphoma', 'Secondary malignant neoplasm of nervous system', 'Malignant neoplasm of eyeball excluding conjunctiva, cornea, retina and choroid', 'Primary malignant neoplasm of eye', "Kaposi's sarcoma of soft tissue", 'Primary malignant neoplasm of soft tissues', 'Malignant neoplasm of auditory tube, middle ear and mastoid air cells', 'Overlapping malignant neoplasm of soft tissues', 'Secondary malignant neoplasm of brain and spinal cord', 'Primary malignant neoplasm of brain', "Hodgkin's disease, lymphocytic depletion of extranodal AND/OR solid organ site", 'Primary malignant neoplasm of occipital lobe', 'Malignant lymphoma', "Diffuse non-Hodgkin's lymphoma, large cell", 'Primary malignant neoplasm of cerebral ventricle', 'Secondary malignant neoplastic disease', 'Primary malignant neoplasm of cerebellum', "Hodgkin's disease, lymphocytic depletion of lymph nodes of head, face AND/OR neck", "Burkitt's tumor of lymph nodes of axilla AND/OR upper limb", 'Primary malignant neoplasm of testis', 'Primary malignant neoplasm of cranial nerve', 'Primary malignant neoplasm of temporal lobe', "Hodgkin's disease, nodular sclerosis of extranodal AND/OR solid organ site", "Kaposi's sarcoma", 'Primary malignant neoplasm of parotid gland', 'B-cell lymphoma', 'Malignant mast cell tumor of intrathoracic lymph nodes', 'Secondary malignant neoplasm of mediastinum', "Hodgkin's disease of lymph nodes of axilla AND/OR upper limb", 'Primary malignant neoplasm of short bone of lower limb', 'Peripheral T-cell lymphoma', "Hodgkin's disease, nodular sclerosis of lymph nodes of head, face and neck", "Hodgkin's disease, mixed cellularity of extranodal AND/OR solid organ site", 'Nodular lymphoma of lymph nodes of head, face and neck', 'Nodular lymphoma of intrathoracic lymph nodes', 'Primary malignant neoplasm of short bone of upper limb', "Hodgkin's paragranuloma of lymph nodes of head, face AND/OR neck", 'Malignant mast cell tumor of lymph nodes of head, face AND/OR neck', 'Malignant lymphoma of intrathoracic lymph nodes', 'Malignant tumor of ascending colon', "Burkitt's tumor of extranodal AND/OR solid organ site", 'Primary malignant neoplasm of undescended testis', 'Multiple myeloma in remission', 'Letterer-Siwe disease of lymph nodes of head, face AND/OR neck', 'Primary malignant neoplasm of long bone of lower limb', "Hodgkin's disease of extranodal AND/OR solid organ site", "Hodgkin's disease of lymph nodes of inguinal region AND/OR lower limb", 'Primary malignant neoplasm of unspecified site', 'Reticulosarcoma', 'Primary malignant neoplasm of cerebral meninges', "Hodgkin's disease, lymphocytic-histiocytic predominance of lymph nodes of head, face and neck", 'Malignant histiocytosis of extranodal AND/OR solid organ site', 'Multiple myeloma', 'Primary malignant neoplasm of labia majora', "Burkitt's tumor of lymph nodes of head, face AND/OR neck", "Hodgkin's sarcoma of extranodal AND/OR solid organ site", "Hodgkin's disease of lymph nodes of head, face AND/OR neck", 'Primary malignant neoplasm of parietal lobe', 'Reticulosarcoma of lymph nodes of head, face and neck', 'Primary malignant neoplasm of nasal cavity', "Hodgkin's disease, lymphocytic-histiocytic predominance of extranodal AND/OR solid organ site", 'Lymphosarcoma of lymph nodes of head, face and neck', 'Essential thrombocythemia', 'Primary malignant neoplasm of mediastinum',

'Malignant lymphoma of lymph nodes of head, face AND/OR neck', 'Disseminated malignancy of unknown primary', "Hodgkin's disease, lymphocytic-histiocytic predominance of lymph nodes of axilla AND/OR upper limb", 'Malignant melanoma of skin of upper limb', "Hodgkin's disease, lymphocytic depletion of lymph nodes of head, face and neck", 'Primary malignant neoplasm', 'Secondary malignant neoplasm of intrathoracic lymph nodes', 'Letterer-Siwe disease of extranodal AND/OR solid organ site', 'Malignant melanoma of skin of neck', "Hodgkin's disease, mixed cellularity of lymph nodes of head, face and neck", 'Malignant lymphoma of extranodal AND/OR solid organ site', 'Secondary malignant neoplasm of lymph nodes of face', "Hodgkin's disease, lymphocytic-histiocytic predominance of intrathoracic lymph nodes", "Hodgkin's granuloma of intrathoracic lymph nodes", 'Malignant neoplasm of tongue, tip and lateral border', "Hodgkin's paragranuloma of extranodal AND/OR solid organ site", "Hodgkin's disease of intrathoracic lymph nodes", 'Primary malignant neoplasm of frontal lobe', 'Reticulosarcoma of lymph nodes of axilla and upper limb', 'Malignant lymphoma of lymph nodes of axilla AND/OR upper limb', 'Malignant histiocytosis of lymph nodes of head, face AND/OR neck', 'Primary malignant neoplasm of brain stem', 'Leukemic reticuloendotheliosis of intrathoracic lymph nodes', 'Primary malignant neoplasm of bronchus', 'Malignant melanoma of skin of scalp', "Hodgkin's paragranuloma of lymph nodes of inguinal region AND/OR lower limb", "Hodgkin's paragranuloma of lymph nodes of axilla AND/OR upper limb", "Hodgkin's granuloma of lymph nodes of head, face AND/OR neck", "Hodgkin's disease, nodular sclerosis of lymph nodes of head, face AND/OR neck", "Hodgkin's disease, mixed cellularity of lymph nodes of inguinal region AND/OR lower limb", "Hodgkin's disease, mixed cellularity of lymph nodes of head, face AND/OR neck", "Hodgkin's disease, lymphocytic-histiocytic predominance of lymph nodes of inguinal region AND/OR lower limb", "Hodgkin's disease, lymphocytic-histiocytic predominance of lymph nodes of head, face AND/OR neck", 'Secondary malignant neoplasm of spleen', 'Secondary malignant neoplasm of lymph nodes of neck', 'Secondary malignant neoplasm of axillary lymph nodes', 'Malignant tumor of thymus', 'Malignant tumor of sigmoid colon', 'Malignant tumor of appendix', 'Malignant tumor of transverse colon', 'Malignant tumor of stomach', 'Malignant tumor of lung', 'Malignant tumor of rectum', 'Malignant tumor of cecum', 'Malignant neoplastic disease', 'Malignant tumor of large intestine', 'Malignant tumor of small intestine', 'Malignant tumor of intestine', 'Malignant tumor of peritoneum', 'Malignant neoplasm of bone', 'Malignant neoplasm of gastrointestinal tract', 'Malignant neoplasm of brain', 'Letterer-Siwe disease of lymph nodes of axilla AND/OR upper limb', 'Clear cell carcinoma of right kidney', 'Peripheral T-cell lymphoma of axillary lymph node', 'Secondary malignant neoplasm of bilateral lungs', 'Myeloid leukemia in relapse', 'Lymphoid leukemia in relapse', 'Liposarcoma of retroperitoneum', 'Renal cell carcinoma of bilateral kidneys', 'Malignant carcinoid tumor of appendix', 'Relapse multiple myeloma', 'Primary extramedullary malignant tumor of spinal cord', 'Liver cell carcinoma', 'Hepatoblastoma', "Diffuse non-Hodgkin's lymphoma, lymphoblastic", 'Primary malignant neoplasm of meninges', 'Gamma heavy chain disease', 'Histiocytic sarcoma', 'Acute promyelocytic leukemia, FAB M3', 'Angiosarcoma of liver', 'T-cell lymphoma', 'Acute panmyelosis with myelofibrosis', 'Refractory anemia with ringed sideroblasts', 'Acute myelomonocytic leukemia, FAB M4', 'Adult T-cell leukemia/lymphoma', 'Overlapping malignant neoplasm of brain', "Diffuse non-Hodgkin's lymphoma", 'Overlapping malignant neoplasm of bone and articular cartilage', "Hodgkin's disease", "Non-Hodgkin's lymphoma", 'Hodgkin lymphoma, nodular lymphocyte predominance', 'Lymphocyte-rich classical Hodgkin lymphoma', "Hodgkin's disease, nodular sclerosis", "Hodgkin's disease, mixed cellularity", 'Hairy cell leukemia', 'Langerhans cell histiocytosis, disseminated', 'Malignant mast cell tumor', 'Mycosis fungoides', 'Malignant histiocytosis', "Hodgkin's disease, lymphocytic depletion", "Burkitt's lymphoma", 'Orbital lymphoma', 'Adenocarcinoma of endometrium', 'Medulloblastoma of cerebellum', 'Malignant white blood cell disorder', 'Follicular low grade B-cell lymphoma', 'Anaplastic astrocytoma of brain', 'T-cell prolymphocytic leukemia', 'Acute megakaryoblastic leukemia', 'High grade B-cell lymphoma', 'Rhabdomyosarcoma of bladder', 'Acute biphenotypic leukemia', 'B-cell chronic lymphocytic leukemia', 'Precursor B-cell acute lymphoblastic leukemia', 'T-cell acute lymphoblastic leukemia', 'Acute monoblastic leukemia', 'Cutaneous/peripheral T-cell lymphoma', 'Malignant teratoma of mediastinum', 'Neuroblastoma of brain', 'Pelvic neuroblastoma', 'Abdominothoracic neuroblastoma', 'Adrenal neuroblastoma', 'Thoracic neuroblastoma', 'Paraspinal neuroblastoma', 'Malignant tumor of lower third of esophagus', 'Malignant tumor of peritoneum and retroperitoneum', 'Malignant neoplasm of pelvic bones, sacrum and coccyx', 'Malignant neoplasm soft tissues of cervical spine', 'Malignant neoplasm of connective and soft tissue of hip and lower limb', 'Malignant neoplasm of connective and soft tissue of perineum', 'Malignant melanoma of axilla', 'Malignant melanoma of breast', 'Malignant neoplasm of upper-outer quadrant of female breast', 'Malignant neoplasm of axillary tail of female breast', 'Malignant neoplasm of cerebrum', 'Malignant tumor of hypothalamus', 'Malignant neoplasm of floor of cerebral ventricle', 'Malignant neoplasm of peripheral nerve of abdomen', "Burkitt's lymphoma of intrathoracic lymph nodes", 'Malignant neoplasm of anterior epiglottis', 'Malignant tumor of thoracic part of esophagus', 'Malignant tumor of body of pancreas', 'Malignant tumor of nasal vestibule', 'Malignant neoplasm of upper lobe of lung', 'Malignant neoplasm of connective and soft tissue of upper limb and shoulder', 'Malignant neoplasm of connective and soft tissue of hand', 'Malignant neoplasm of connective and soft tissue of thorax', 'Malignant neoplasm of connective and soft tissue of abdominal wall', 'Malignant melanoma of auricle', 'Malignant melanoma of hip', 'Malignant melanoma of thigh', 'Malignant neoplasm of scalp and/or skin of neck', 'Malignant neoplasm of central part of female breast', 'Malignant neoplasm of upper-inner quadrant of female breast', 'Choriocarcinoma', 'Malignant tumor of undescended testis', 'Malignant tumor of lacrimal gland', 'Malignant tumor of choroid plexus', 'Malignant neoplasm of peripheral nerves and autonomic nervous system', 'Malignant neoplasm of peripheral nerve of thorax', 'Diffuse malignant lymphoma - small non-cleaved cell', 'Adult T-cell leukemia', 'Malignant tumor of posterior wall of oropharynx', 'Malignant tumor of posterior wall of nasopharynx', 'Malignant neoplasm of calcaneum', 'Malignant neoplasm of connective and soft tissue of finger', 'Malignant neoplasm of connective and soft tissue of lower leg', 'Malignant neoplasm of connective and soft tissue of abdomen', 'Malignant melanoma of scalp', 'Malignant tumor of ectopic testis', 'Malignant neoplasm, overlapping lesion of bladder', 'Malignant neoplasm of globus pallidus', 'Malignant neoplasm of pons', 'Malignant tumor of cranial nerve', 'Malignant neoplasm of pituitary gland and craniopharyngeal duct', 'Malignant tumor of nasopharynx', 'Malignant tumor of anterior wall of nasopharynx', 'Malignant tumor of abdominal part of esophagus', 'Malignant tumor of body of stomach', 'Malignant neoplasm of rectum, rectosigmoid junction and anus', 'Malignant neoplasm of liver and intrahepatic bile ducts', 'Malignant tumor of Islets of Langerhans', 'Malignant neoplasm of ribs and/or sternum and/or clavicle', 'Malignant neoplasm of first metatarsal bone', 'Malignant neoplasm of third metatarsal bone', 'Malignant melanoma of scalp and/or neck', 'Malignant melanoma of neck', 'Malignant melanoma of upper arm', 'Malignant melanoma of ankle', 'Malignant neoplasm of skin of ear and external auditory canal', 'Malignant neoplasm of lower-inner quadrant of female breast', 'Malignant neoplasm of endometrium of corpus uteri', 'Malignant tumor of trigone of urinary bladder', 'Malignant tumor of bladder neck', 'Malignant neoplasm of peripheral nerves of head, face and neck', 'Compound leukemias', 'Malignant histiocytosis of lymph nodes of inguinal region and lower limb', 'Malignant neoplasm of peripheral nerve of pelvis', "Burkitt's lymphoma of intra-abdominal lymph nodes", 'Chloroma', 'Fibrolamellar hepatocellular carcinoma', 'Merkel cell carcinoma', 'Peripheral neuroectodermal tumor', 'Neuroendocrine carcinoma', 'Small cell carcinoma of lung', 'Acral lentiginous malignant melanoma of skin', 'Cutaneous fibrosarcoma', 'Familial cancer of breast', 'Endodermal sinus tumor of ovary', 'Clear cell carcinoma of kidney', 'Astrocytoma of brain', 'Rhabdomyosarcoma of orbit', 'Malignant melanoma of iris', 'Follicular thyroid carcinoma', 'Medullary thyroid carcinoma', 'Squamous cell carcinoma of skin of cheek', 'IgG myeloma', 'Squamous cell carcinoma of skin', 'Metastasis to lymph node of unknown primary', 'Adenocarcinoma of lung', 'Basal cell carcinoma of skin', 'Metastasis to lung of unknown primary', 'Metastasis to adrenal gland of unknown primary', 'Malignant tumor of breast', 'Malignant phyllodes tumor of breast', 'Mucinous cystadenocarcinoma of ovary', 'Granulosa cell tumor of ovary', 'Malignant germ cell tumor of ovary', 'Sarcoma of kidney', 'Oligodendroglioma of brain', 'Astrocytoma of spinal cord', 'Carcinoma of bladder', 'Pituitary carcinoma', 'Malignant tumor of unknown origin', 'Malignant tumor of head and neck', 'Carcinoma of parotid gland', 'Sarcoma of liver', 'Malignant epithelial tumor of ovary', 'Dysgerminoma of ovary', 'Carcinoma of prostate', 'Rhabdomyosarcoma', 'Synovial sarcoma', 'Malignant melanoma of choroid', 'Papillary thyroid carcinoma', 'Adrenal carcinoma', 'Parathyroid carcinoma', 'Malignant tumor of salivary gland', 'Malignant tumor of digestive organ', 'Malignant tumor of soft tissue of head, face and neck', 'Malignant lymphoma - small lymphocytic', 'Nephroblastoma', 'Carcinoma of lower limb bones', 'Carcinoma of genital organ', 'Aleukemic leukemia', 'Metastasis to head and neck lymph node', 'Chronic myelomonocytic leukemia', 'Osteosclerotic myeloma', 'Philadelphia chromosome-positive acute lymphoblastic leukemia', 'Precursor B-cell acute lymphoblastic leukemia in remission', 'Acute myeloid leukemia with maturation, FAB M2, in remission', 'Erythroleukemia, FAB M6 in remission', 'Retroperitoneal sarcoma', 'Acute myelomonocytic leukemia, FAB M4, in remission', 'Malignant lymphoma in remission', 'Monosomy 7 syndrome', 'T-cell acute lymphoblastic leukemia in remission', "Ewing's sarcoma of bone", "Hodgkin's disease in remission", 'Solitary osseous myeloma', 'Metastatic adenocarcinoma of unknown origin', 'Osteosarcoma of bone', 'Malignant tumor of lymphoid hemopoietic and related tissue', "Follicular non-Hodgkin's lymphoma", 'Nodular lymphoma', 'Malignant melanoma of head and neck', 'Malignant melanoma of upper limb', 'Malignant melanoma of back', 'Malignant tumor of lower labial mucosa', 'Malignant neoplasm of upper lobe, bronchus or lung', 'Malignant melanoma of lower limb', 'Secondary nodes - axilla/arm', 'Malignant neoplasm of canthus', 'Acute lymphoblastic leukemia, transitional pre-B-cell', 'Malignant neoplasm of soft tissue', 'Malignant melanoma of trunk', 'Malignant neoplasm of genitourinary organ', 'Malignant neoplasm of central nervous system', 'Malignant neoplasm of skin', 'Malignant neoplasm of body of uterus', 'Malignant tumor of soft tissue of upper limb', 'Malignant neoplasm of nervous system', 'Malignant neoplasm of female breast', 'Malignant neoplasm of skin of trunk', 'Secondary malignant neoplasm of unknown site', 'Retinoblastoma', 'Adenocarcinoma of prostate', 'Adamantinoma of femur', 'Malignant neoplasm of adrenal cortex', 'Primary malignant neoplasm of vocal cord', 'Malignant neoplasm of bone of lower limb', 'Primary malignant neoplasm of breast', 'Malignant melanoma', 'Malignant neoplasm of skin of lower limb', 'Malignant neoplasm of skin of upper limb', 'Malignant neoplasm of upper limb bones and scapula', 'Malignant tumor of prostate', 'Metastasis to digestive organs', 'Palate carcinoma', 'Malignant tumor of urinary system', 'Malignant melanoma of eye', 'Malignant glioma of spinal cord', 'B-cell acute lymphoblastic leukemia', 'Low grade B-cell lymphoma', 'B-cell prolymphocytic leukemia', 'Dermatofibrosarcoma protuberans', 'Lymphoma of intestine', 'Malignant glioma of brain', 'Glioblastoma multiforme of brain', 'Squamous cell carcinoma of tongue', 'Malignant tumor of lateral wall of nasopharynx', 'Malignant tumor of extrahepatic bile duct', 'Malignant tumor of nasal cavity', 'Malignant tumor of ethmoid sinus', 'Malignant tumor of sphenoid sinus', 'Malignant tumor of vertebral column', 'Malignant tumor of vagina', 'Malignant tumor of testis', 'Malignant tumor of urethra', 'Malignant tumor of conjunctiva', 'Malignant neoplasm of occipital lobe', 'Malignant neoplasm of cerebral ventricles', 'Malignant tumor of spinal cord', 'Malignant tumor of pituitary gland', 'Malignant tumor of pelvis', 'Malignant tumor of meninges', 'Malignant tumor of lower limb', 'Malignant tumor of pharynx', 'Malignant tumor of female genital organ', 'Malignant tumor of vulva', 'Malignant tumor of vermilion border of lower lip', 'Malignant tumor of tongue', 'Malignant tumor of gum', 'Malignant tumor of palate', 'Malignant tumor of ampulla of Vater', 'Malignant tumor of head of pancreas', 'Malignant tumor of larynx', 'Malignant neoplasm of labia majora', 'Malignant tumor of scrotum', 'Malignant tumor of ureter', 'Malignant tumor of retina', 'Malignant neoplasm of cerebral meninges', 'Malignant tumor of thyroid gland', 'Malignant tumor of pineal gland', 'Malignant tumor of neck', 'Primary malignant neoplasm of gastrointestinal tract', 'Malignant basal cell neoplasm of skin', 'Malignant tumor of lip', 'Malignant tumor of anal canal', 'Malignant tumor of parotid gland', 'Malignant tumor of sublingual gland', 'Malignant tumor of buccal mucosa', 'Malignant tumor of hard palate', 'Malignant tumor of colon', 'Malignant tumor of pancreas', 'Malignant retroperitoneal tumor', 'Malignant tumor of pleura', 'Malignant tumor of heart', 'Malignant tumor of orbit', 'Malignant neoplasm of temporal lobe', 'Malignant neoplasm of brainstem', 'Malignant tumor of vocal cord', 'Malignant tumor of optic nerve', 'Malignant tumor of gallbladder', 'Malignant tumor of adrenal gland', 'Malignant tumor of soft tissue of hip', 'Malignant tumor of base of tongue', 'Malignant tumor of major salivary gland', 'Malignant tumor of soft palate', 'Malignant tumor of uvula', 'Malignant tumor of oropharynx', 'Malignant tumor of tonsil', 'Malignant tumor of esophagus', 'Malignant tumor of splenic flexure', 'Malignant tumor of biliary tract', 'Malignant tumor of maxillary sinus', 'Malignant tumor of frontal sinus', 'Malignant tumor of ovary', 'Malignant tumor of foreskin', 'Malignant tumor of epididymis', 'Malignant tumor of renal pelvis', 'Malignant neoplasm of eye', 'Malignant tumor of choroid', 'Malignant neoplasm of frontal lobe', 'Malignant neoplasm of parietal lobe', 'Malignant neoplasm of spinal meninges', 'Malignant tumor of parathyroid gland', 'Malignant tumor of mediastinum', 'Malignant tumor of upper limb', 'Malignant tumor of nasal sinuses', 'Malignant tumor of male genital organ', 'Smoldering myeloma', 'Refractory cytopenia with multilineage dysplasia', 'Metastatic neuroblastoma of orbit proper', 'Acute monocytic/monoblastic leukemia', 'Metastasis from malignant tumor of bone', 'Leukemic infiltrate of retina', 'Malignant neoplasm of lower eyelid', 'Local recurrence of malignant tumor of lung', 'Metastasis from malignant tumor of breast', 'Local recurrence of malignant tumor of thyroid gland', 'Local recurrence of malignant tumor of breast', 'Metastasis from malignant tumor of kidney', 'Metastasis from malignant tumor of lung', 'Metastasis from malignant tumor of thyroid', 'Primary osteosarcoma of pelvis', 'Extranodal NK/T-cell lymphoma, nasal type', 'Malignant sacral teratoma', 'Plasmacytoma', 'Malignant teratoma of testis', 'Gestational choriocarcinoma', "Kaposi's sarcoma associated with AIDS", 'Precursor T-cell lymphoblastic lymphoma', 'Malignant optic glioma', 'Widespread metastatic malignant neoplastic disease', 'Infiltrating duct carcinoma of breast', 'Malignant melanoma of skin of cheek', 'Malignant melanoma of skin of ankle', 'Primary malignant neoplasm of forearm', 'Malignant neoplasm of liver', 'Primary malignant neoplasm of optic nerve', 'Secondary malignant neoplasm of bone marrow', 'Secondary malignant neoplasm of brain', 'Primary malignant neoplasm of blood vessel', 'Primary malignant neoplasm of bone marrow', 'Primary malignant neoplasm of central nervous system', 'Primary cutaneous T-cell lymphoma', 'Gorlin syndrome', 'Basal cell carcinoma of nose', 'Basal cell carcinoma of cheek', 'Squamous cell carcinoma of skin of ear', 'Dermatofibrosarcoma protuberans with granular cell change', 'Spindle cell rhabdomyosarcoma', 'Low-grade fibromyxoid sarcoma', 'Primary cutaneous marginal zone B-cell lymphoma', 'Basal cell carcinoma of upper extremity', 'Congenital malignant melanoma', 'Squamous cell carcinoma of foot', 'Squamous cell carcinoma of skin of lower extremity', 'Basal cell carcinoma of scalp', 'Epithelioid hemangioendothelioma', 'Lymphangiosarcoma', 'Alveolar rhabdomyosarcoma', 'Extrarenal rhabdoid tumor', 'Malignant melanoma of soft tissues', 'Squamous cell carcinoma', 'Squamous cell carcinoma of anogenital area', 'Squamous cell carcinoma of scalp', 'Squamous cell carcinoma of forehead', 'Infantile fibrosarcoma', 'Malignant Triton tumor', 'Embryonal rhabdomyosarcoma', 'Myxoid liposarcoma', 'Pleomorphic liposarcoma', 'Yolk sac tumor', 'Precursor B-cell lymphoblastic lymphoma involving skin', 'Malignant peripheral nerve sheath tumor', 'Botryoid rhabdomyosarcoma', 'Alveolar soft part sarcoma', 'Leukemic infiltration of skin', 'Olfactory neuroblastoma', 'Adenoid cystic carcinoma of submandibular gland', 'Rhabdomyosarcoma of connective or soft tissue', 'Mucoepidermoid carcinoma of parotid gland', 'Malignant glioma of brainstem', 'Sarcoma of head and neck', 'Malignant tumor of spinal cord, extramedullary', 'Primary sarcoma of tongue', 'Mucoepidermoid carcinoma of submandibular gland', 'Malignant neoplasm of skin of eyelid', 'Mucoepidermoid carcinoma of salivary gland', 'Sarcoma', 'Primary malignant neoplasm of respiratory tract', 'Secondary malignant neoplasm of back', 'Secondary malignant neoplasm of chest wall', 'Secondary malignant neoplasm of bone of lower limb', 'Secondary malignant neoplasm of cerebral meninges', 'Secondary malignant neoplasm of orbit', 'Malignant tumor of spinal cord, intramedullary', 'Primary malignant neoplasm of sacrococcygeal region', 'Primary malignant neoplasm of skin of lower limb', 'Primary malignant neoplasm of trunk', 'Secondary malignant neoplasm of bladder', 'Secondary malignant neoplasm of head', 'Adenoid cystic carcinoma of parotid gland', 'Secondary malignant neoplasm of pelvis', 'Sarcoma of soft tissue', 'Anaplastic glioma of brain', 'Secondary malignant neoplasm of vertebral column', 'Malignant neoplasm of thoracic cavity structure', 'Neuroblastoma', 'Malignant neoplasm of lower respiratory tract', 'Basal cell carcinoma of eyelid', 'Squamous cell carcinoma of eyelid', 'Renal medullary carcinoma', 'Pineoblastoma', 'Adenosarcoma of cervix uteri', 'Acquired cystic disease associated renal cell carcinoma', 'Differentiation syndrome due to and following chemotherapy co-occurrent with acute promyelocytic leukemia', 'Acute myeloid leukemia with inv(3)(q21q26.2) or t(3;3)(q21;q26.2); RPN1-EVI1', 'Low grade astrocytoma of brain', 'Oligodendroglioma of cerebrum', 'Peripheral T-cell lymphoma of lymph nodes of multiple sites', 'Small lymphocytic B-cell lymphoma of intra-abdominal lymph nodes', "Non-Hodgkin's lymphoma of lymph nodes of multiple sites", 'Malignant germ cell neoplasm of right ovary', 'Malignant germ cell neoplasm of left ovary', 'Malignant melanoma of left choroid', 'Malignant melanoma of right choroid', 'Acute lymphoid leukemia relapse', 'Small cell neuroendocrine carcinoma of bladder', 'Refractory thrombocytopenia', 'Acute leukemia of ambiguous lineage', 'Systemic Epstein-Barr virus positive T-cell lymphoproliferative disease of childhood', 'Primary synovial sarcoma of soft tissue of limb', 'Primary malignant neuroendocrine neoplasm of appendix', 'Primary rhabdomyosarcoma of pharynx', 'Primary malignant neuroendocrine neoplasm of lung', 'Malignant epithelial neoplasm', 'Primary rhabdomyosarcoma of male genital organ', 'Primary embryonal carcinoma of testis', 'Astrocytoma of brain stem', 'Non-seminomatous germ cell neoplasm of testis', 'Ganglioneuroblastoma', 'Primary malignant astrocytoma of central nervous system', 'Malignant neoplasm of chest wall', 'Prostate cancer metastatic to bone', 'Malignant germ cell tumor of testis', 'Philadelphia chromosome-negative precursor B-cell acute lymphoblastic leukemia', 'Kaposi sarcoma of viscus', 'Ewing sarcoma of bone of pelvis', 'Anaplastic astrocytoma of central nervous system', 'Malignant glioma of cerebrum', 'Malignant glioma of hypothalamus', 'Malignant glioma of central nervous system', 'Ewing sarcoma of bone structure of upper limb', 'Astrocytoma of cerebrum', 'Malignant poorly differentiated neuroendocrine carcinoma', 'Disorder of central nervous system co-occurrent and due to acute lymphoid leukemia', 'Malignant neuroendocrine tumor', 'Primary malignant neoplasm of right kidney', 'Malignant neoplasm of lower lobe of right lung', 'Malignant neoplasm of upper lobe of left lung', 'Papillary renal cell carcinoma', 'Nodal marginal zone B-cell lymphoma', 'Epithelioid sarcoma', 'Primary malignant neoplasm of left adrenal gland', "Refractory Hodgkin's lymphoma", 'Nephroblastoma of left kidney', 'Nephroblastoma of right kidney', 'Malignant immature teratoma of ovary', 'Juvenile myelomonocytic leukemia in remission', 'B-cell prolymphocytic leukemia in remission',

'Interdigitating dendritic cell sarcoma', 'Ependymoblastoma', 'Medulloepithelioma', 'Pineal parenchymal tumor of intermediate differentiation', 'ALK-positive large B-cell lymphoma', 'Embryonal sarcoma of liver', 'Squamous cell carcinoma of head and neck', 'Malignant neoplasm of dorsal surface of tongue', 'Pulmonary blastoma', 'Malignant teratoma', 'Malignant neoplasm of skin of face', 'Metastatic sarcoma', 'Malignant fibromatous neoplasm', 'Medulloblastoma', 'Malignant fibrous histiocytoma', 'Primary mediastinal (thymic) large B-cell lymphoma', 'Acute myeloid leukemia with t(9:11)(p22;q23); MLLT3-MLL', 'Mantle cell lymphoma', 'Metastatic malignant melanoma', 'Chondrosarcoma', 'Malignant neoplasm of ear', 'Seminoma', 'Malignant neoplasm of skeletal system', 'Leiomyosarcoma', 'Juvenile myelomonocytic leukemia', 'Malignant carcinoid tumor', 'Extranodal marginal zone B-cell lymphoma of mucosa-associated lymphoid tissue', 'Oligodendroglioma', 'Mixed glioma', 'Malignant adenomatous neoplasm', 'Leiomyosarcoma of scalp', 'Malignant germ cell neoplasm of anterior mediastinum', 'Sarcoma of tibia', 'Diffuse follicle center lymphoma', 'Malignant neoplasm of anterior mediastinum', 'Nasopharyngeal carcinoma', 'Sarcoma of mandible', 'Carcinoma of female breast', 'Chondrosarcoma of bone', "Non-Hodgkin's lymphoma of tonsil", 'Sarcoma of scapula', 'Sarcoma of skull', 'Sarcoma of pelvis', 'Malignant neoplasm of cerebellum', "Ewing's sarcoma of soft tissue", "Non-Hodgkin's lymphoma of extranodal site", 'Anaplastic large cell lymphoma, ALK negative', 'Carcinoma of thyroid', 'Malignant neoplasm of cerebellopontine angle', 'Malignant neoplasm of urinary organ', 'Malignant neoplasm of connective tissue', 'Sarcoma of bone of foot', 'Chordoma of clivus', 'Malignant teratoma of retroperitoneum', 'Marginal zone lymphoma', 'Malignant neoplasm of mandible', 'Malignant neoplasm of posterior mediastinum', 'Sarcoma of bone', 'Sarcoma of femur', 'Carcinoma of pineal gland', 'Malignant neoplasm of intraabdominal organ', 'Squamous cell carcinoma of nose', 'Carcinoma of brain', 'Carcinoma of lung', "Follicular non-Hodgkin's lymphoma of tonsil", 'Malignant neoplasm of respiratory system', 'Glioblastoma multiforme', 'Pediatric follicular lymphoma', 'Astroblastoma of brain', 'Classical Hodgkin lymphoma', 'Malignant neoplasm of long bone of lower limb', 'Malignant neoplasm of long bone of lower leg', 'Primary adenocarcinoma of pancreas', 'Metastasis to lymph node from adenocarcinoma', 'Supratentorial primitive neuroectodermal tumor', 'Primitive neuroectodermal tumor', 'Paratesticular malignant neoplasm', 'Sarcoma of orbit', 'Endometrial stromal sarcoma', 'Renal cell carcinoma', 'Primary myoepithelial carcinoma of oropharynx', 'Pleuropulmonary blastoma', 'Pleuropulmonary blastoma type III', 'Therapy-related myelodysplastic syndrome', 'Metastatic neuroblastoma', 'Metastatic renal cell carcinoma', 'Secondary malignant neoplasm of right lung', 'Diffuse sclerosing papillary thyroid carcinoma', 'Malignant neoplasm of superior wall of nasopharynx', 'Malignant neoplasm of short bone of lower limb', 'Secondary malignant neoplasm of left lung'

**Disease included under Sickle cell disease:**

'Sickle cells [Presence] in Blood by Light microscopy', 'Sickle cell anemia in mother complicating childbirth', 'Sickle cell retinopathy', 'Sickle cell-hemoglobin SS disease', 'Sickle cell-beta-thalassemia', 'Sickle cell beta plus thalassemia', 'Sickle cell-beta^0^-thalassemia', 'Sickle cell-delta beta^0^-thalassemia', 'Sickle cell anemia with coexistent alpha-thalassemia', 'Sickle cell-hemoglobin Lepore disease', 'Acute chest syndrome due to sickle cell-hemoglobin C disease with crisis', 'Proliferative retinopathy due to sickle cell disease', 'Sickle cell-hemoglobin C retinopathy', 'Sickle cell anemia with high hemoglobin F', 'Sickle cell-hemoglobin D disease', 'Sickle cell-hemoglobin C disease', 'Sickle cell-thalassemia disease', 'Priapism due to sickle cell disease', 'Hand-foot syndrome in sickle cell anemia', 'Hemoglobin SS disease without crisis', 'Sickle cell-hemoglobin D disease without crisis', 'Sickle cell-hemoglobin E disease with crisis', 'Sickle cell-hemoglobin E disease without crisis', 'Sickle cell-thalassemia disease with crisis', 'Sickle cell-thalassemia disease without crisis', 'Hemoglobin SS disease with crisis', 'Sickle cell-hemoglobin C disease with crisis', 'Sickle cell-hemoglobin C disease without crisis', 'Sickle cell-hemoglobin D disease with crisis', 'Hemoglobin SS disease with vasoocclusive crisis', 'History of sickle cell anemia', 'Acute sickle cell splenic sequestration crisis', 'Sickle cell-hemoglobin E disease', 'Renal papillary necrosis due to sickle cell disease', 'Focal segmental glomerulosclerosis due to sickle cell disease', 'Vaso-occlusive pain episode in sickle cell disease', 'HBB (hemoglobin, subunit beta) (eg, sickle cell anemia, beta thalassemia, hemoglobinopathy); duplication/deletion variant(s)', 'HBB (hemoglobin, subunit beta) (eg, sickle cell anemia, beta thalassemia, hemoglobinopathy); full gene sequence'
